# Pre- and postnatal environmental lithium exposure and adolescent neurodevelopment: a tooth-biomarker study

**DOI:** 10.64898/2026.11.06.26367460

**Authors:** Maximilian König, Andrew T. Marshall, Shana Adise, Eric Kan, Rob McConnell, Mauro Martinez, Vishal Midya, Manish Arora, Elizabeth R. Sowell

**Author notes:** Corresponding Author: Elizabeth R. Sowell, PhD Department of Pediatrics, Division of Neurology, Keck School of Medicine Children’s Hospital Los Angeles, University of Southern California 4650 Sunset Blvd., Mailstop #130, Los Angeles, CA 90027 |. These authors contributed equally to this work (Maximilian König, Andrew T. Marshall).

## Abstract

Lithium is a naturally occurring element found in drinking water, and its environmental presence is rising with demand for batteries. Yet evidence on lithium comes almost entirely from adults, ecological studies, or medical treatment, leaving the effects of prenatal and early postnatal environmental exposure unknown. We laser-mapped lithium in shed baby teeth, which preserve a record of exposure around birth (-12 to +13 weeks), from 448 U.S. youth in the Adolescent Brain Cognitive Development Study and tested associations with brain structure, cognition, and behavior at ages 9-10. Higher lithium was found in children of less-educated caregivers and from more deprived neighborhoods. Higher lithium was also linked to thicker cortices, varying surface area, mostly smaller subcortical volumes, poorer reading and language ability, and more externalizing problems, with associations varying by caregiver education.

Early-life environmental lithium exposure may therefore be socioeconomically patterned and contribute to disparities in neurodevelopment, warranting environmental health surveillance.

## Introduction

Lithium is a naturally occurring trace element with psychiatric importance and expanding environmental relevance. Beyond its role as a first-line mood stabilizer for bipolar disorder (*1*, *2*), lithium is central to rechargeable battery technologies (*3*) and is present in soils, groundwater, and drinking water at concentrations that vary widely by region but are not federally regulated in the United States (*4*).

Ecological studies, which compare average exposure and outcomes across communities rather than individuals, have linked higher naturally occurring drinking-water lithium to lower suicide mortality (*5*, *6*), whereas other work has raised concerns, including associations with autism spectrum disorder (*7*) and altered thyroid function (*8*). Critically, this evidence derives almost entirely from adult populations, ecological designs, or therapeutic exposure. Lithium supplementation of drinking water for suicide prevention is actively debated (*9*, *10*). Yet, the neurodevelopmental implications of low-level early-life environmental lithium exposure remain poorly understood.

The prenatal and early postnatal periods are sensitive windows during which exposures can lastingly affect neural structure, cognition, and behavior (*11*, *12*). Lithium crosses the placenta and blood-brain barrier, enabling fetal and early postnatal brain exposure (*13*, *14*), and can influence intracellular signaling, neurotrophic processes (which support the survival and growth of neurons), synaptic plasticity (the strengthening and weakening of connections between neurons), and gray matter structure (*15*, *16*), making atypical brain development a biologically plausible outcome. Adolescence is an informative period for detecting the imprint of such exposure, because it reflects both earlier neurodevelopmental processes and the ongoing maturation of systems supporting cognition, emotion, and behavior (*17*).

Another question is who is exposed. Socioeconomic context may shape both the likelihood of lithium exposure, through residential location, water source, and housing infrastructure, and vulnerability to its effects (*18*, *19*). County-level data suggest that U.S. areas with higher groundwater lithium have lower average income and education (*20*), but whether an individual child’s early-life lithium exposure is socioeconomically patterned has not been examined. If it is, lithium could be an overlooked environmental contributor to disparities in child development.

Shed deciduous (“baby”) teeth offer a rare opportunity to address these gaps. Teeth begin mineralizing prenatally, and trace elements are incorporated into dentine, the tooth’s inner layer, in incremental growth layers, much like tree rings, that preserve a temporally ordered record of exposure. A visible marker of birth within the tooth, the neonatal line, allows these layers to be aligned to birth and resolved to specific weeks of development (*21*, *22*). This captures internalized lithium exposure at a developmental timescale that drinking-water concentrations, residential estimates, and later-childhood biospecimens such as blood cannot.

Using dentine biomarkers from participants in the Adolescent Brain Cognitive Development (ABCD) Study, a large multisite U.S. cohort that enrolled children at ages 9 to 10 (*23*), we examined whether prenatal and early postnatal environmental lithium exposure was associated with adolescent brain structure, cognition, and behavior. We measured brain structure with magnetic resonance imaging (MRI), focusing on cortical thickness (the depth of the brain’s outer layer), cortical surface area (its extent), and subcortical volume (the size of gray matter structures below the cerebral cortex, such as the hippocampus and amygdala); cognition with the NIH Toolbox; and emotional and behavioral problems with the Child Behavior Checklist, including internalizing problems (e.g., anxiety and depression) and externalizing problems (e.g., aggression and rule-breaking). Given limited and indirect prior evidence, we made no regional or directional predictions for brain or cognitive outcomes, though lithium’s mood-stabilizing role led us to explore whether lithium exposure was associated with fewer internalizing problems. We further asked whether lithium exposure was patterned by socioeconomic context, and whether socioeconomic context modified its neurodevelopmental associations, considering three indicators: household income, caregiver education, and geocoded measures of neighborhood deprivation.

## Results

### Early-life lithium was reconstructed from baby teeth in 448 adolescents

Of 4,708 youth aged 9 to 10 years in the ABCD Study who donated at least one shed deciduous tooth, 500 teeth (one per participant) underwent dentine metal analysis, 448 (89.6%) of which met quality- control criteria and were analyzed (226 [50.4%] female; mean [SD] age, 9.94 [0.64] years; Table 1).

**Table 1:**
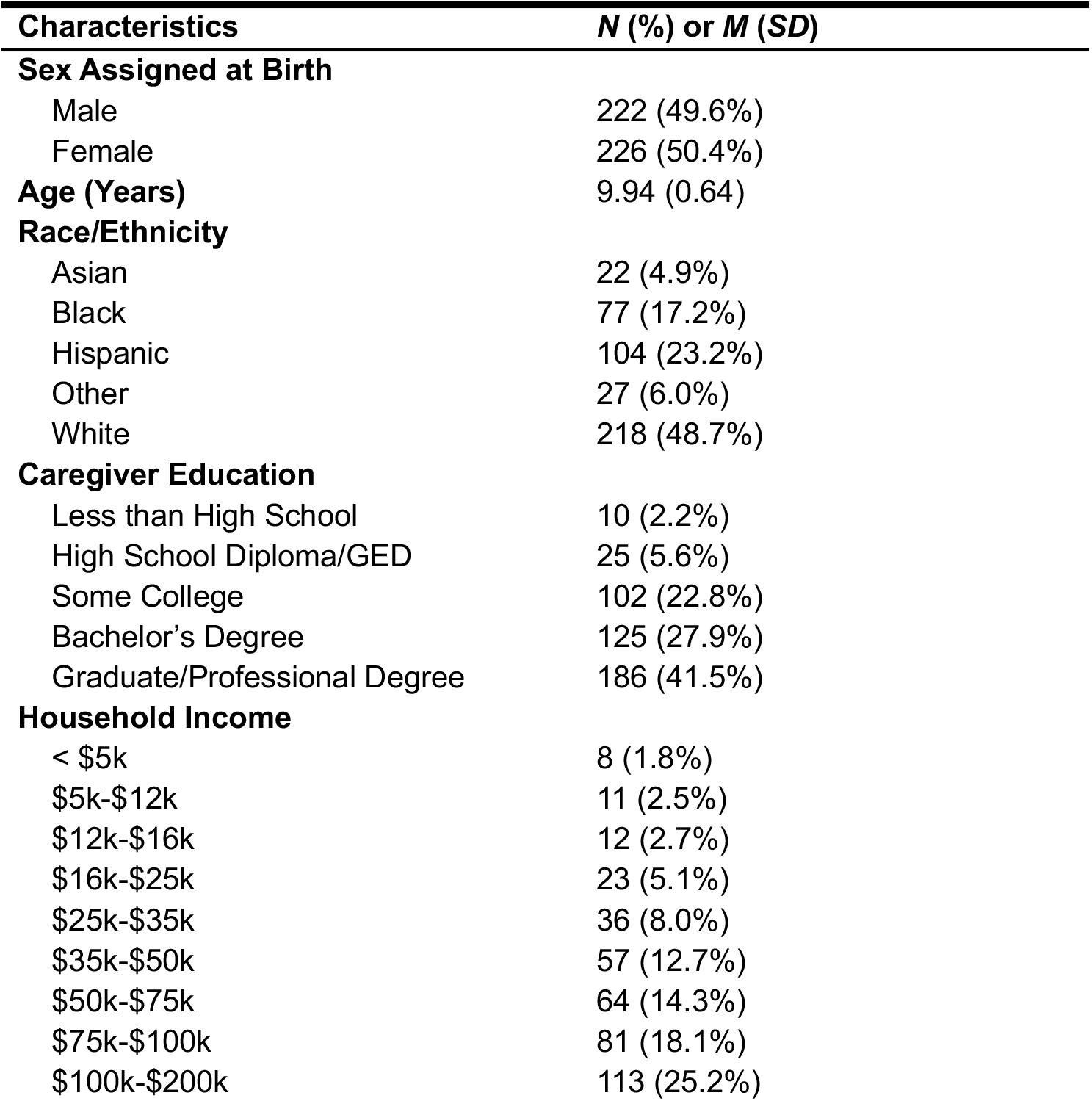
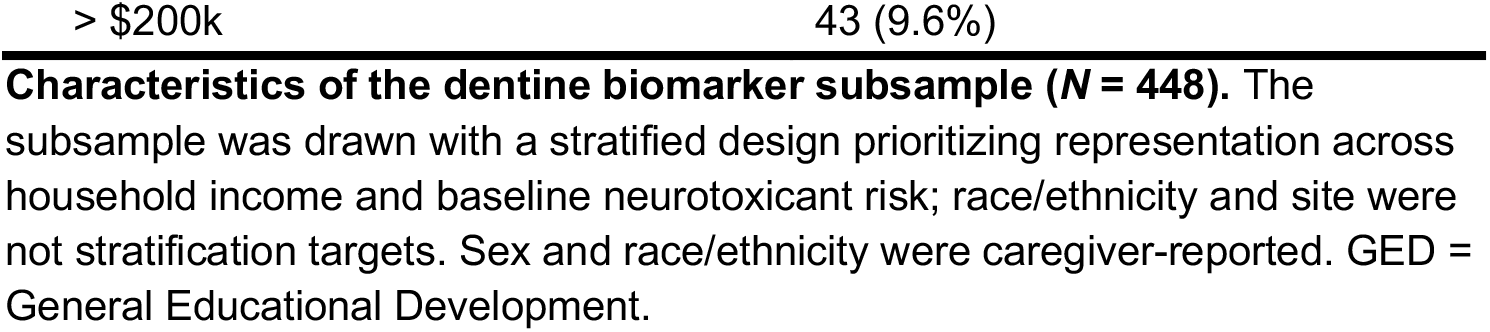
Characteristics of the dentine biomarker subsample (*N* = 448). The subsample was drawn with a stratified design prioritizing representation across household income and baseline neurotoxicant risk; race/ethnicity and site were not stratification targets. Sex and race/ethnicity were caregiver-reported. GED = General Educational Development.

| **Characteristics** | ***N* (%) or *M* (*SD*)** |
|---|---|
| **Sex Assigned at Birth** |  |
| Male | 222 (49.6%) |
| Female | 226 (50.4%) |
| **Age (Years)** | 9.94 (0.64) |
| **Race/Ethnicity** |  |
| Asian | 22 (4.9%) |
| Black | 77 (17.2%) |
| Hispanic | 104 (23.2%) |
| Other | 27 (6.0%) |
| White | 218 (48.7%) |
| **Caregiver Education** |  |
| Less than High School | 10 (2.2%) |
| High School Diploma/GED | 25 (5.6%) |
| Some College | 102 (22.8%) |
| Bachelor’s Degree | 125 (27.9%) |
| Graduate/Professional Degree | 186 (41.5%) |
| **Household Income** |  |
| < \$5k | 8 (1.8%) |
| \$5k-\$12k | 11 (2.5%) |
| \$12k-\$16k | 12 (2.7%) |
| \$16k-\$25k | 23 (5.1%) |
| \$25k-\$35k | 36 (8.0%) |
| \$35k-\$50k | 57 (12.7%) |
| \$50k-\$75k | 64 (14.3%) |
| \$75k-\$100k | 81 (18.1%) |
| \$100k-\$200k | 113 (25.2%) |
| > \$200k | 43 (9.6%) |

Dentine lithium (measured relative to calcium, as the ^7^Li:^43^Ca ratio) spanning 12 weeks before to 13 weeks after birth was quantified by laser ablation-inductively coupled plasma mass spectrometry (LA-ICP- MS), along growth layers aligned to birth using the neonatal line (Table S1, Fig. S1-S2). Dentine lithium was then related to socioeconomic context, brain structure (cortical thickness, surface area, and subcortical volume), cognition (NIH Toolbox), and emotional-behavioral problems (Child Behavior Checklist). All outcomes were analyzed using linear mixed-effects models and reported as partial correlations (*rp*) with false discovery rate correction for multiple testing.

### Lithium exposure was patterned by caregiver education and neighborhood deprivation

Higher lithium was associated with lower caregiver education (*rp* = -.16, 95% CI [-.25, -.07], *p* < .001) and greater neighborhood deprivation (*rp* = .12, 95% CI [.01, .23], *p* = .036), but not household income (*rp* = -.07, *p* = .151) (Tables S2.1–S4.2). Because caregiver education showed the strongest association with dentine lithium it was retained as the primary socioeconomic covariate for the remaining analyses.

### Lithium was associated with regionally specific differences in brain structure

Across imaging modalities, lithium exposure showed structural associations that differed in regional specificity (Fig. 1; Fig. S3; supplementary results). For cortical thickness, 41 of 68 regions showed significant positive associations, with the strongest effect in the right caudal middle frontal gyrus (*rp* = .11, 95% CI [.09, .13], *p* < .001) (Fig. 1A; Fig. S3A; Tables S5.1–S5.2). For cortical surface area, 45 of 68 regions were significant, with bidirectional effects (strongest positive: left isthmus cingulate, *rp* = .13, 95% CI [.11, .15], *p* < .001; strongest negative: right superior frontal gyrus, *r*p = -.10, 95% CI [-.11, -.08], p < .001) (Fig. 1B; Fig. S3B; Tables S6.1–S6.2). For subcortical volume, 10 of 16 regions were significant and effects were predominantly smaller (strongest: right accumbens, *rp* = -.12, 95% CI [-.14, -.10], *p* < .001), with positive associations limited to the bilateral hippocampus (right: *rp* = .05, 95% CI [.03, .07], *p* < .001; left: *rp* = .04, 95% CI [.02, .05], *p* < .001) (Fig. 1C; Fig. S3C; Tables S7.1–S7.2).

**Fig. 1.**
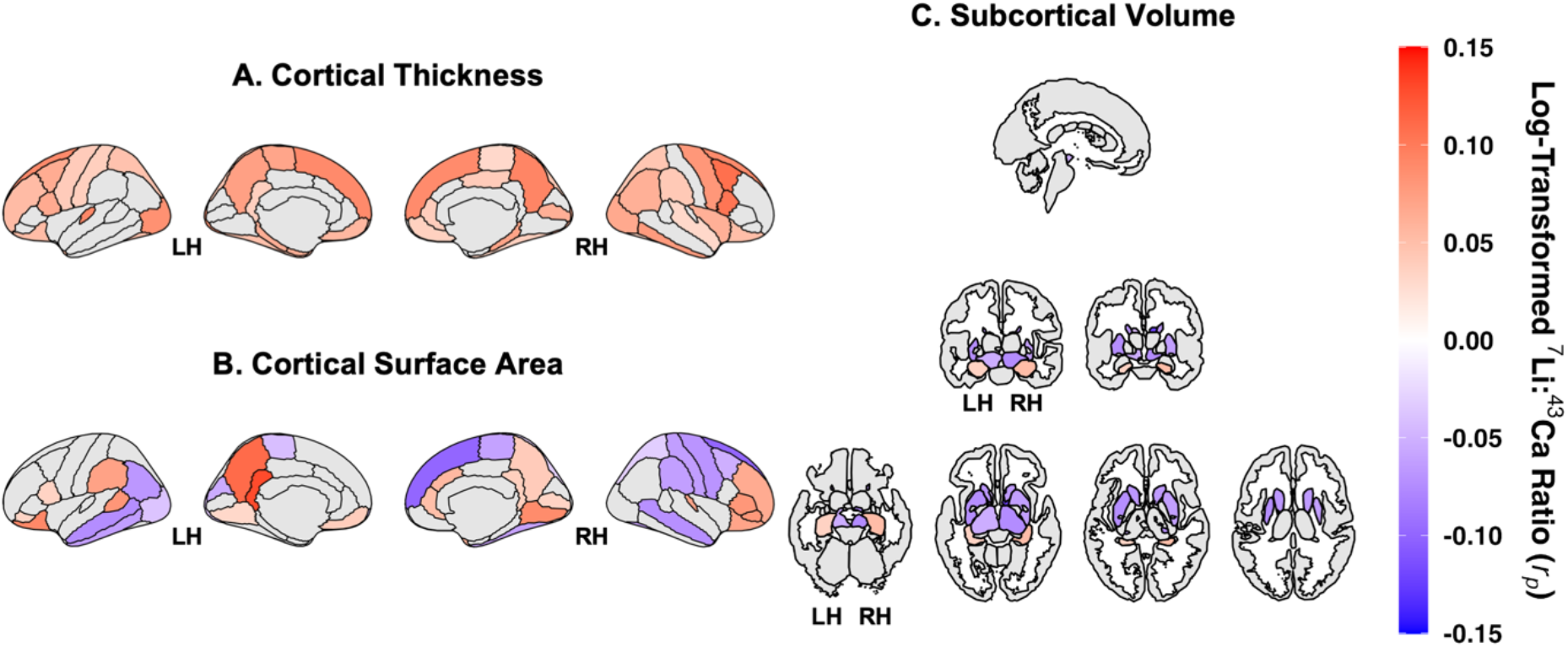
Whole-brain associations between dentine lithium levels and adolescent brain structure. Surface-based and subcortical maps display partial correlations (*rp*) between participants’ mean log- transformed ^7^Li:^43^Ca ratio and (**A**) cortical thickness, (**B**) cortical surface area, and (**C**) subcortical volume. Cortical maps are shown for the left and right hemisphere (LH, RH) in lateral and medial views.

Subcortical structures are visualized across multiple axial and coronal slices. Color values represent partial correlation coefficients (*rp*) derived from covariate-adjusted linear mixed-effects models, including age, sex, weeks since birth (WSB) (-12 to +13), caregiver education, and modality-specific global brain measures. Warmer colors indicate positive associations, and cooler colors indicate negative associations. Only regions surviving false discovery rate (FDR) correction (Benjamini-Hochberg, α = .005) are displayed; non-significant regions are shown in gray. Analyses were conducted after exclusion of influential observations identified via Cook’s distance (99^th^ percentile threshold; *n* = 5 excluded), yielding a final sample of *N* = 432. The number of total observations per region ranged from 11,120 to 11,126 due to repeated lithium measurements across timepoints.

### Caregiver education moderated the brain associations

Caregiver education moderated these neuroanatomical associations (Fig. 2; Fig. S4). For cortical thickness, the positive associations between dentine lithium levels and cortical thickness were generally stronger among youth of more-educated caregivers, with 28 of 68 lithium **×** caregiver-education interactions significant (71% positive) and the strongest interaction in the left rostral anterior cingulate (*rp* = .13, 95% CI [.11, .15], *p* < .001) (Fig. 2A; Table S5.3). For surface area, associations between dentine lithium levels and surface area varied heterogeneously as a function of caregiver education, with 44 of 68 interactions significant (50% positive) and the strongest interaction in the left precentral gyrus (*rp* = -.16, 95% CI [-.17, -.14], *p* < .001) (Fig. 2B; Table S6.3). For subcortical volume, caregiver education moderated lithium’s associations in a region-specific manner: the already-positive association between lithium and hippocampal volume was strengthened among youth of more-educated caregivers, while negative associations in regions such as the caudate and ventral diencephalon were attenuated. Eight of 16 interactions were significant (100% positive), with the strongest interaction in the right hippocampus (*rp* = .10, 95% CI [.08, .12], *p* < .001) (Fig. 2C; Table S7.3).

**Fig. 2.**
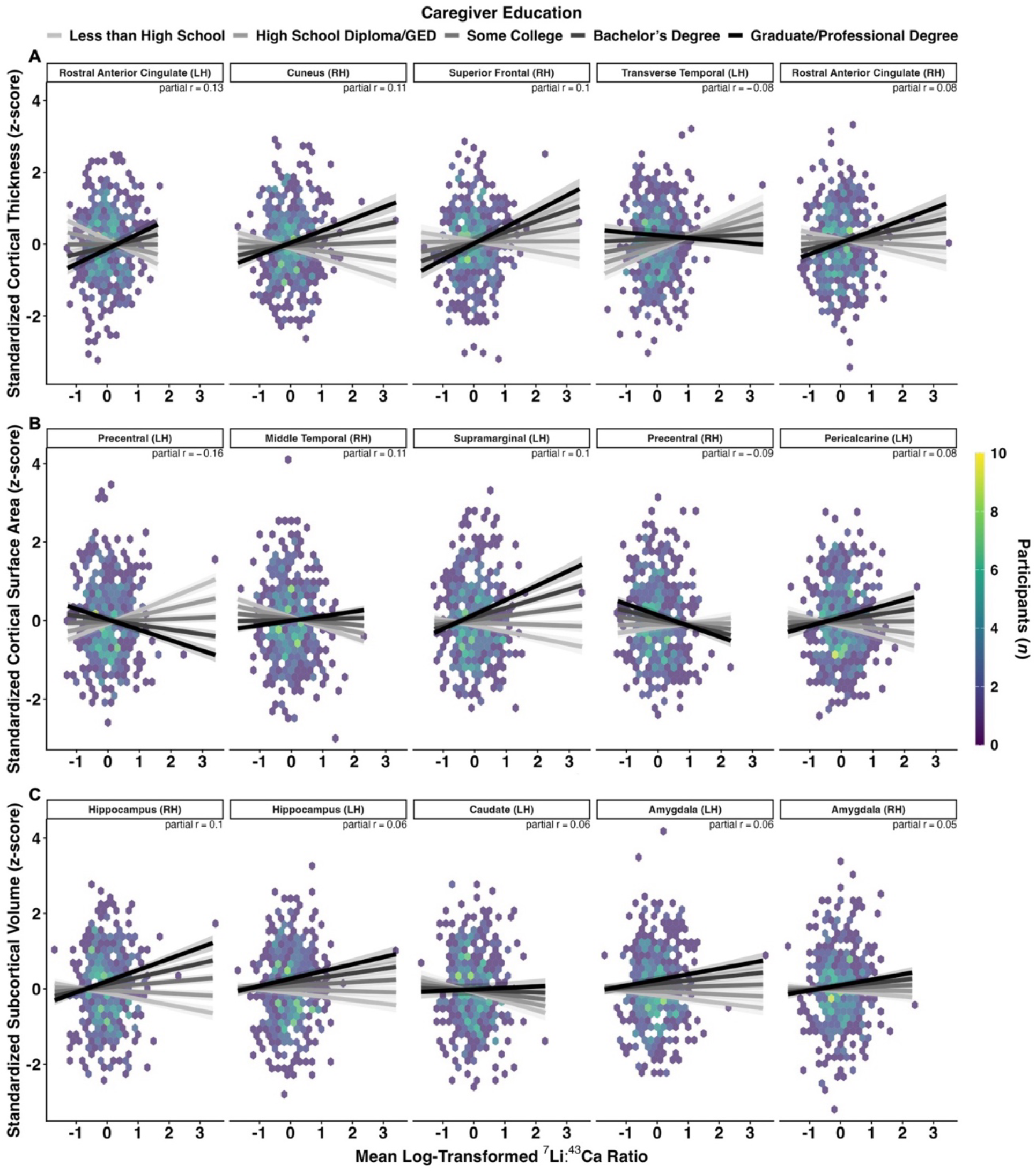
Caregiver education moderated associations between dentine lithium levels and adolescent brain structure. Hexbin plots display associations between participants’ mean log-transformed ^7^Li:^43^Ca ratio (x-axis) and standardized (**A**) cortical thickness, (**B**) cortical surface area, and (**C**) subcortical volume measures (*z*-scores; y-axis) across regions showing the strongest lithium **×** caregiver education interactions following FDR correction (Benjamini-Hochberg, α = .005). Regions were ranked by partial correlations (*rp*). Each hexagon represents the number of participants within bins of early-life environmental lithium exposure and brain structure measures, with brighter colors indicating greater density. Lines represent model-implied regression fits stratified by caregiver education level, ranging from less than high school to graduate/professional degree, with shaded bands indicating 95% confidence intervals (delta method), following exclusion of influential observations identified using Cook’s distance (99^th^ percentile threshold; *n* = 5 excluded). Linear mixed-effects models were adjusted for age, sex, weeks since birth (-12 to +13), caregiver education, and modality-specific global brain measures, including whole brain volume for cortical thickness analyses, total cortical surface area for cortical surface area analyses, and intracranial volume for subcortical volume analyses. The analytic sample after exclusion was *N* = 432, with the number of observations per region ranging from 11,120 to 11,126 due to repeated lithium measurements across timepoints. LH = left hemisphere; RH = right hemisphere.

### Lithium was linked to poorer reading, buffered by caregiver education

Higher lithium was associated with lower total cognition (*rp* = -.03, 95% CI [-.05, -.01], *p* = .008), driven by the crystallized composite (*rp* = -.06, 95% CI [-.08, -.04], *p* < .001) and specifically Oral Reading Recognition (*rp* = -.14, 95% CI [-.16, -.11], *p* < .001; a 1.27-point decrease per interquartile-range increase in dentine lithium, 95% CI [-1.44, -1.09]). Fluid cognition was not associated with lithium (*rp* = .001, *p* = .892). Caregiver education positively moderated several cognitive associations, including Oral Reading Recognition (*rp* = .13, 95% CI [.11, .16], *p* < .001), such that the negative association between lithium and Oral Reading Recognition was attenuated to a near-null association among youth of more-educated caregivers (Fig. 3A; Fig. S5, S7A; Tables S8.1–S8.3, S11, S12; supplementary results).

**Fig. 3.**
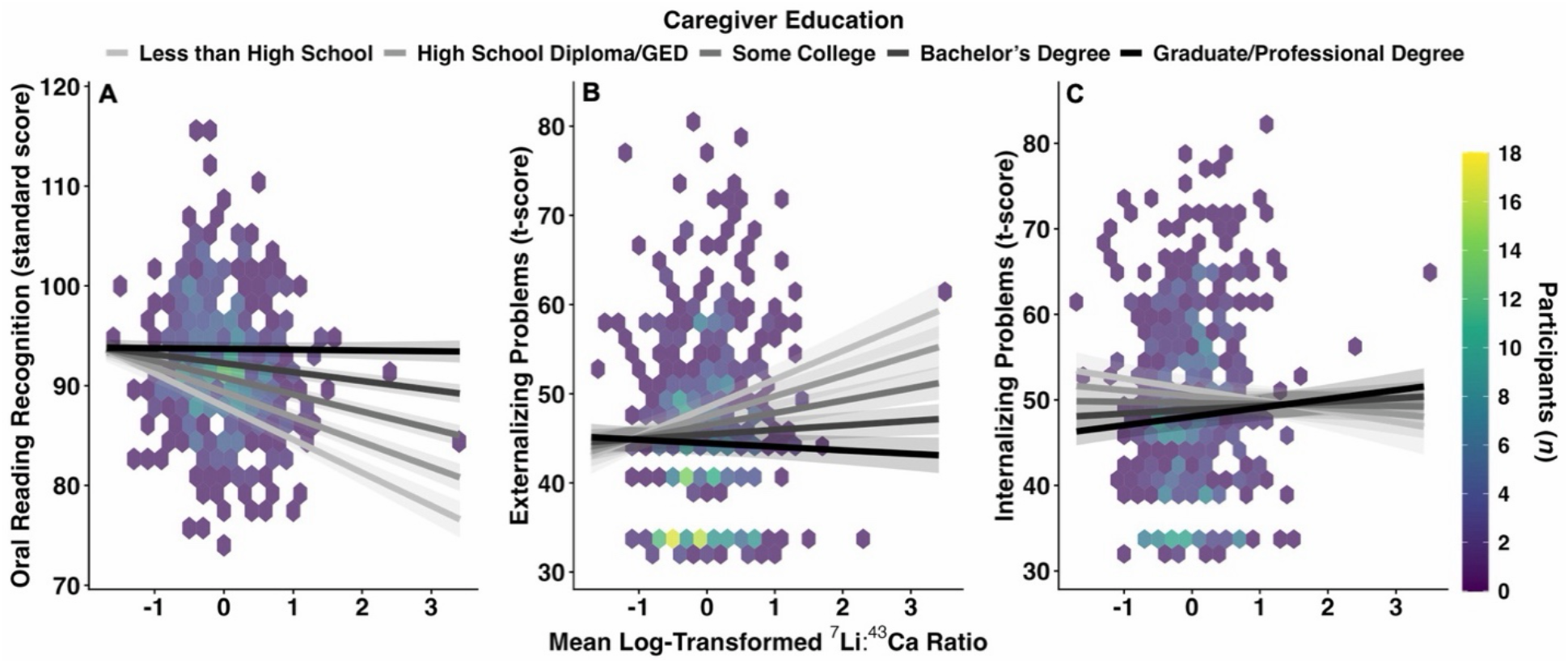
Caregiver education moderated associations between dentine lithium levels and cognitive and emotional-behavioral outcomes. Hexbin plots display associations between participants’ mean log- transformed ^7^Li:^43^Ca ratio (x-axis) and (**A**) Oral Reading Recognition (standard score), (**B**) externalizing problems (*t*-score), and (**C**) internalizing problems (*t*-score). All plotted lithium **×** caregiver interactions survived FDR correction (Benjamini-Hochberg, α = .05). Colored hexagons represent the number of participants within bins of early-life environmental lithium exposure and outcome measures, with brighter colors indicating greater density. Lines represent model-implied regression fits stratified by caregiver education level, ranging from less than high school to graduate/professional degree, with shaded bands indicating 95% confidence intervals (delta method), following exclusion of influential observations identified using Cook’s distance (99^th^ percentile threshold; *n* = 5 excluded). Linear mixed-effects models were adjusted for age, sex, weeks since birth (-12 to +13), and caregiver education. The analytic sample after exclusion was *N* = 439 for panel A, *N* = 443 for panel B, and *N* = 443 for panel C. The number of observations per outcome ranged from 11,303 to 11,406 due to repeated lithium measurements across timepoints.

### Lithium was linked to externalizing problems, moderated by caregiver education

Higher lithium was associated with greater total problems (*rp* = .05, 95% CI [.03, .07], *p* < .001) and externalizing problems (*rp* = .05, 95% CI [.03, .07], *p* < .001; a 1.39-point increase per interquartile-range increase in dentine lithium, 95% CI [1.11, 1.67]) but not internalizing problems (*rp* = .02, *p* = .082).

Caregiver education attenuated the externalizing association (*rp* = -.08, 95% CI [-.10, -.06], *p* < .001) (Fig. 3B) but positively moderated the internalizing association (*rp* = .05, 95% CI [.03, .07], *p* < .001) (Fig. 3C), indicating that higher lithium was linked to more internalizing problems specifically among youth of more- educated caregivers. This pattern, together with the externalizing findings, suggests that caregiver education does not uniformly buffer against lithium’s behavioral associations (Fig. S6, S7B; Tables S9.1– S9.3, S11, S12; supplementary results).

### Lithium-associated brain regions were unrelated to cognition and behavior

In follow-up analyses, none of the lithium-associated brain regions were associated with crystallized cognition, fluid cognition, or internalizing and externalizing problems (Table S10), arguing against a structural mediation pathway.

## Discussion

In a demographically diverse developmental cohort, prenatal and early postnatal environmental lithium exposure, reconstructed from analyses of shed baby teeth, was socioeconomically patterned and associated with adolescent brain structure, cognition, and behavior. Environmental lithium exposure was higher among youth with less-educated caregivers and in more deprived neighborhoods, extending environmental-justice concerns to a naturally occurring, unregulated element. The same dentine exposure tracked with different neurodevelopmental correlates depending on social context.

Cortical thickness was the most robust structural correlate, with higher lithium associated with thicker cortex across frontal, parietal, and temporal regions. This direction is consistent with reports of greater cortical thickness, reduced cortical thinning, or thickness values closer to healthy controls among lithium- treated patients with bipolar disorder (*24–27*). Although therapeutic and environmental exposures differ in dose and timing, this convergence suggest that lithium’s association with cortical structure may extent to lower, environmental exposure levels.

Cortical surface area showed a less uniform pattern, with higher lithium associated with larger and smaller surface area across frontal, parietal, temporal, occipital, and cingulate cortices. This bidirectional pattern does not contradict the thickness findings, as thickness and surface area are partly distinct features of cortical organization with different genetic and developmental determinants (*28*). Early-life environmental lithium exposure thus appears associated with regional variation in cortical arealization. Unlike the positive surface-area effects reported in ENIGMA’s lithium-treated bipolar cohort, including a larger left paracentral lobule (*27*), the bidirectional pattern suggests therapeutic lithium findings may not generalize to early environmental exposure.

Subcortical volumes of the striatum and diencephalon were predominantly smaller with higher lithium exposure. The bilateral hippocampus was a notable exception, showing positive associations. The latter echoes neuroprotective hippocampal findings in the adult clinical lithium literature (*29*, *30*), whereas the smaller striatal and diencephalic volumes do not fit a uniform model. Together, the structural findings indicate regionally specific associations rather than a single global pattern. However, these structural associations did not extend to cognitive or behavioral outcomes, arguing against structural mediation.

Associations with cognition and behavior were domain-specific: lower Oral Reading Recognition and more externalizing problems, without an internalizing association. No comparable population-based literature has quantified real-world neurodevelopmental effect sizes for environmental lithium exposure, so whether these associations are clinically large or small remains unclear. For context, our effect sizes resemble those reported for lead, a well-characterized pediatric neurotoxicant: each doubling of dentine lithium was associated with a 1.18-point decrement in Oral Reading Recognition and a 1.31-point increase in externalizing problems, versus a 1.84-point IQ decrement (*31*) and a 2.15-point increase in externalizing problems (*32*) per doubling of blood lead in prior cohorts, though these comparisons span different biomarkers, cohorts, and outcomes and warrant caution.

These outcome domains differ from prior adolescent environmental-lithium studies, which reported inverse associations between drinking-water lithium and depressive symptoms, interpersonal violence, or psychotic experiences (*33*, *34*). The one overlapping domain, depressive symptoms, is addressed by our internalizing measure. The absence of an internalizing association does not support a generalized mood- related benefit of early-life environmental lithium and cautions against extrapolating from adult ecological studies linking drinking-water lithium to lower suicide rates (*5*, *6*). These results align with a mixed developmental literature, including null findings for attention-deficit/hyperactivity disorder (*35*) and for neuropsychological functioning after prenatal therapeutic lithium (*36*), alongside a reported association with autism spectrum disorder (*7*).

The socioeconomic patterning of environmental lithium exposure is the most consequential result. Higher dentine lithium among youth from less advantaged households and neighborhoods aligns with evidence that unequal exposure burdens extend beyond industrial contaminants to naturally occurring constituents in drinking water (*37–39*). Unlike area-level drinking-water estimates used by most prior studies, dentine biomarkers reconstruct week-specific, individual-level lithium exposure prospectively linked to outcomes assessed nearly a decade later. This approach is among the closest approximations to a gold-standard lithium-epidemiology design. The dentine approach shows that reconstructed early-life environmental lithium exposure is itself socioeconomically patterned. Dentine biomarkers cannot identify lithium exposure sources, such as drinking-water source and groundwater reliance, diet, or housing plumbing and water-treatment infrastructure, and future work should link these biomarkers to residential and water- source data. This effort is important as lithium demand expands for electric-vehicle batteries and grid- scale storage, which may reshape environmental exposure pathways and their social/geographical distribution (*40*).

Caregiver education predicted lithium exposure and moderated its associations with neurodevelopment across brain, cognition, and behavior. For brain structure, higher lithium was associated with greater cortical thickness and larger subcortical volumes among youth with higher caregiver education, while surface area showed positive and negative interactions across regions. For cognition, caregiver education buffered the association between higher lithium and poorer reading decoding/language ability to a near- null level among youth with more-educated caregivers. Behavioral moderation was less uniform: higher caregiver education was associated with a buffered externalizing association but an elevated internalizing association, underscoring caregiver education’s mixed protective and deleterious effects. This moderation likely reflects context dependence rather than a biological effect of education itself. Caregiver education indexes many co-varying features of the developmental environment, including cognitive stimulation, nutrition, co-exposures, healthcare access, and residential stability. These factors, rather than education per se, likely explain the observed moderation. They may act by altering toxicant absorption or neurodevelopmental vulnerability (*41–43*), though the biological mechanisms remain speculative and warrant preclinical follow-up.

These findings are relevant to active debates about supplementing drinking water with lithium for suicide prevention (*9*, *10*). Our study was not designed to evaluate that policy, and the results should not be read as evidence for or against it. However, they indicate that policy-relevant evidence cannot be limited to adult psychiatric outcomes and must weigh early-life, neurodevelopmental, trace-element mixtures, and social-contextual dimensions (*44*).

This study has several limitations. The observational, cross-sectional design precludes causal inference, and residual confounding remains possible despite adjustment. The subsample was selected from tooth donors and for destructive analysis, and it skewed toward higher caregiver education, limiting generalizability, especially to families with fewer educational resources. Dentine biomarkers index internalized lithium exposure but not its source. The analytic window, 12 weeks before to 13 weeks after birth, maximized coverage across participants but limits inference about earlier prenatal or later postnatal exposure. Lithium was examined in isolation, whereas trace elements co-occur and may act jointly.

Behavioral outcomes were caregiver-reported. Effect sizes were small across many regional tests (|*r*p| mostly .03–.16), a magnitude consistent with other single-predictor associations in pediatric neuroimaging, including dentine lead exposure in the same subsample (*45*) and family income and caregiver education with cortical structure (*41*, *46*, *47*). Such small effects are expected given the multifactorial determinants of brain structure and do not preclude population-level relevance amid the socioeconomic patterning of exposure. Independent replication and longitudinal follow-up will be essential.

In conclusion, early-life environmental lithium exposure was developmentally relevant and socioeconomically patterned. Its downstream correlates were further moderated by caregiver education. The findings do not establish environmental lithium exposure as beneficial or harmful but suggest that developmental studies of environmental lithium should be interpreted separately from adult and therapeutic research, and that early-life and social-contextual evidence should inform any consideration of population-level exposure strategies.

## Materials and Methods

### Study design and participants

This cross-sectional analysis used baseline data from the ABCD Study (7.0 data release; doi:10.82525/8f3w-5260), a prospective cohort that enrolled 11,878 youth aged 9 to 10 years across 21 U.S. sites between 2016 and 2018 using school-based sampling (*23*). A centralized institutional review board at the University of California, San Diego, provided ethical oversight, with approvals obtained at each site. Caregivers provided written informed consent and youth provided assent. Reporting follows the Strengthening the Reporting of Observational Studies in Epidemiology guideline.

Among 4,708 ABCD participants who donated shed deciduous teeth, 500 were selected for dentine biomarker analysis using a design stratified on household income and baseline neurotoxicant risk to balance socioeconomic and environmental-contaminant risk and limit selection bias versus convenience sampling (*45*). Because the laser ablation-inductively coupled plasma mass spectrometry (LA-ICP-MS) ablates dentine, only a subset was analyzed to preserve the repository for future work. Teeth with caries, restorations, developmental defects, or structural loss were excluded. Of the 500 selected participants, 52 failed biospecimen quality control, leaving 448 (89.6%) as the analytic sample (Table 1; Table S1; Fig. S1) (*45*). Subsample size was based on assay cost and tooth-donor availability rather than a prospective power calculation.

### Dentine lithium

Early-life environmental lithium exposure was quantified along dentine growth layers using LA-ICP-MS (Agilent 8900 ICP-MS with extended-range detection coupled to a New Wave Research NWR-193 [ESI, USA] laser ablation system equipped with a 193-nm ArF excimer laser) (*21*, *22*, *48–52*), with the neonatal line aligning measurements to weeks since birth (WSB) (Fig. 4A) (*21*, *22*). Tooth sections were analyzed for trace elements at 35-μm spatial resolution, yielding approximately 50 developmental time points per tooth and reconstructing weekly exposure profiles spanning late gestation through early postnatal life (Fig. 4B). Lithium was quantified as the calcium-normalized isotope ratio ^7^Li:^43^Ca and log-transformed (*22*). Helium was used as the carrier gas within the laser ablation cell and mixed with argon via a T-piece prior to introduction into the ICP-MS. Instrument performance was optimized daily using NIST SRM 612 standards to maximize sensitivity (maximize ion counts), minimize oxide formation (^232^Th^16^O^+^/^232^Th^+^, < 0.3%), and monitor elemental fractionation (^232^Th^+^/^238^U^+^, 100±5%). Elemental intensities were normalized to calcium to account for variation in mineral density within and across teeth and were externally calibrated using NIST SRM 610, 612, 614 standards. Because tooth types mineralize on different schedules (*53*), participant coverage varied across weeks. The primary analytic window was restricted to 12 weeks before through 13 weeks after birth, when at least 90.8% of the 500 participants contributed data (Fig. S2).

**Fig. 4.**
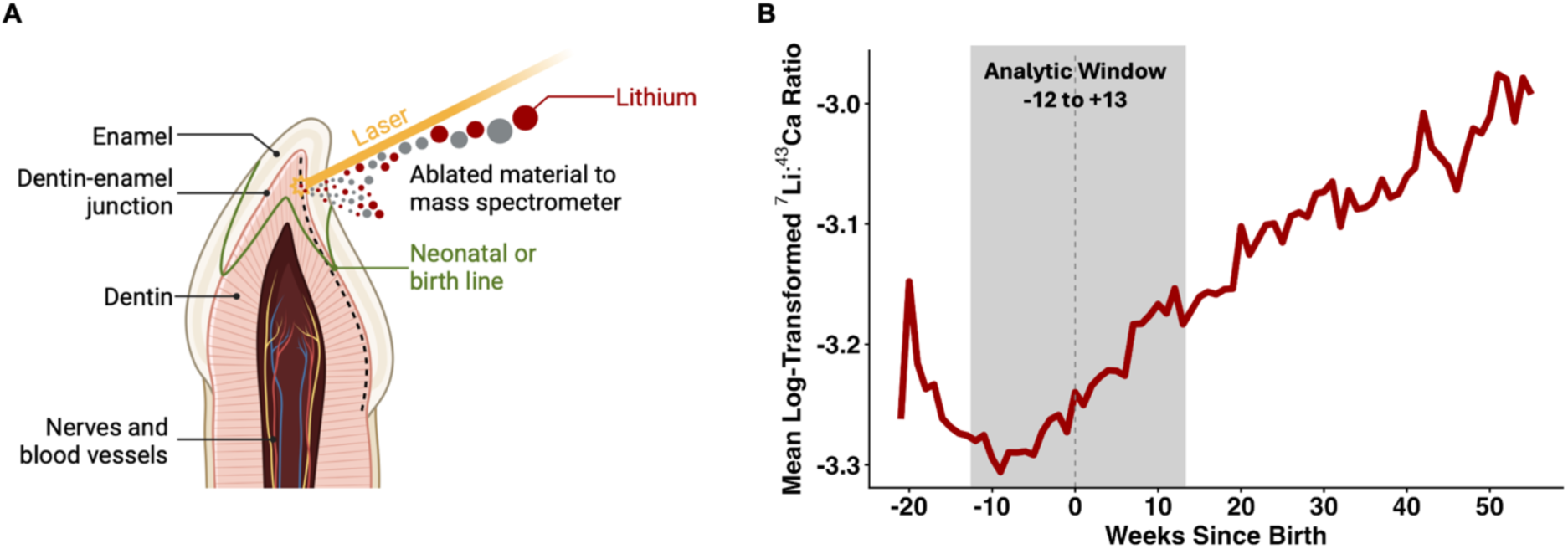
Reconstruction of early-life lithium exposure from deciduous teeth. (**A**) Schematic illustration of laser ablation-inductively coupled plasma mass spectrometry (LA-ICP-MS) sampling along the dentine- enamel junction of a shed deciduous tooth (dashed outline), adapted from Frye and colleagues (*54*). Incremental dentine formation preserves temporally ordered records of trace element incorporation across development. The neonatal line was used as a histological marker of birth to align dentine measurements to developmental timing in weeks since birth (WSB), with negative values representing prenatal periods and positive values representing postnatal periods. Laser-ablated dentine material was analyzed to quantify lithium concentrations relative to calcium (^7^Li:^43^Ca). (**B**) Cohort-level developmental trajectory of mean log-transformed ^7^Li:^43^Ca ratios across prenatal and early postnatal development. The dashed vertical line indicates birth (0 WSB), and the analytic window (-12 to +13 WSB) is shaded in gray. Fig. 4A was created with BioRender.com.

### Brain, cognitive, and behavioral measures

Baseline T1-weighted structural MRI was acquired with harmonized 3T protocols and centrally processed using a uniform, automated FreeSurfer (v7.1.1) pipeline applied across participants regardless of exposure status, limiting differential measurement bias (*55*). Cortical thickness and surface area were derived using FreeSurfer-based cortical reconstruction and parcellation into 68 Desikan-Killiany regions, and subcortical volumes from automated segmentation into 16 regions (*56–58*). Modality-specific global measures (whole-brain volume, total cortical surface area, and intracranial volume) served as covariates. Analyses were restricted to participants passing T1 quality control (*n* = 437 [97.5%]). Additional details regarding ABCD neuroimaging acquisition, preprocessing, and quality control procedures are described elsewhere (*55*).

Cognition was assessed with the NIH Toolbox (*59*), a standardized and validated computerized assessment designed to measure multiple domains of neurocognitive functioning in children and adolescents. The battery consists of tasks assessing crystallized cognition, including Picture Vocabulary (language/vocabulary comprehension) and Oral Reading Recognition (language/reading decoding), as well as fluid cognition domains, including Pattern Comparison Processing Speed (processing speed), Picture Sequence Memory (episodic memory), List Sorting Working Memory (working memory), Flanker Inhibition Control and Attention Test (executive function/attention/inhibition), and Dimensional Change Card Sort (executive function/cognitive flexibility). Composite and subtest scores were used to characterize both global and domain-specific cognitive performance.

Emotional-behavioral problems were assessed with the caregiver-reported Child Behavior Checklist (CBCL) (*60*), a standardized 113-item questionnaire designed to assess emotional and behavioral functioning over the previous six months. Caregivers rated each item using a three-point scale ranging from 0 (not true) to 2 (very true/often true). Standardized scores for internalizing problems, externalizing problems, and total problems were derived, with internalizing and externalizing symptoms used as the primary behavioral outcomes in the present study.

## Statistical analyses

All statistical analyses were conducted in R version 4.5.2 (*61*), primarily using the lme4, lmerTest, effectsize, influence.ME, ggplot2, and ggseg packages, among others, for linear mixed-effects modeling, effect-size estimation, influential-case diagnostics, and visualization. Additional packages were used for data processing, table generation, and figure creation.

Lithium values were log-transformed to reduce skewness and approximate normality. Observations with lithium values ≤ 0 or missing (0.03% of values) were excluded prior to transformation. The resulting log- transformed ^7^Li:^43^Ca ratio served as the primary exposure metric in all analyses. Continuous predictors were mean-centered before analysis, and sex was effect coded (male = -1, female = +1).

Baseline demographic and socioeconomic covariates were obtained from ABCD Study caregiver-report and linked external data measures. Caregivers reported child sex assigned at birth, race/ethnicity, annual household income, and caregiver educational attainment as part of the baseline demographic assessment. Neighborhood socioeconomic context was characterized using the Area Deprivation Index (ADI), available through ABCD Linked External Data, which links geocoded residential information to census-tract-level contextual measures. ADI was used as an indicator of neighborhood socioeconomic disadvantage, capturing broader neighborhood-level contextual features such as poverty, housing quality, employment, and community resources (*62*, *63*), with higher values reflecting greater deprivation.

Census-tract-level identifiers are not released by ABCD to protect participant confidentiality, so tract-level random effects could not be specified in any model; only aggregate tract-derived measures such as ADI are available to researchers. Household income captures material resources available within the home, including access to housing, nutrition, health care, and environmental protections (*18*). Caregiver education reflects caregiver educational attainment and related dimensions of health literacy, occupational opportunity, and developmental enrichment that are not reducible to income (*18*). Because these measures index distinct but overlapping dimensions of socioeconomic environment, associations between early-life environmental lithium exposure and adolescent development may differ depending on whether socioeconomic context is defined at the household, caregiver, or neighborhood level. Pairwise associations among the three indicators were evaluated as partial correlations from linear mixed-effects models with a random intercept for assessment site, the finest clustering structure available given the tract-identifier restriction noted above. These indicators were intercorrelated (household income ∼ caregiver education: *rp* = .57, 95% CI [.51, .63], *p* < .001; household income ∼ neighborhood deprivation: *rp* = -.28, 95% CI [-.36, -.19], *p* < .001; caregiver education ∼ neighborhood deprivation: *rp* = -.36, 95% CI [-.44, -.28], *p* < .001), consistent with related but nonredundant dimensions of socioeconomic context (*18*). Caregiver education showed the strongest association with dentine lithium in preliminary models and was retained as the primary socioeconomic covariate.

Household income was modeled as a continuous midpoint-coded variable derived from the 10-level household income categories (<$5k, $5k-$12k, $12k-$16k, $16k-$25k, $25k-$35k, $35k-$50k, $50k-$75k, $75k-$100k, $100k-$200k, >$200k), using bracket midpoint estimates in thousands of dollars. Caregiver education was modeled using a 5-level ordinal variable: Less than High School, High School Diploma/GED, Some College, Bachelor’s Degree, Graduate/Professional Degree. ADI was modeled as a neighborhood-level measure of socioeconomic disadvantage.

Linear mixed-effects models were used to characterize temporal variation in dentine lithium levels across WSB and to examine associations between dentine lithium levels and neurodevelopmental outcomes.

Missing data were handled via complete-case analysis, excluding rather than imputing participants missing a variable, so analytic *n* varied across models. Initial models examined associations between dentine lithium levels and socioeconomic status (SES) indicators while adjusting for sex and WSB. These models included random intercepts for participants and ABCD assessment site and followed the general form:

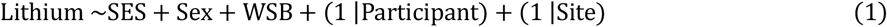

Additional linear-mixed effects models evaluated whether tooth type, attrition severity, and resorption severity should be included as covariates in subsequent analyses. These models examined associations with dentine lithium levels while including fixed effects for sex, WSB, as well as random intercepts for participants and assessment site. Tooth type (*F*(4, 438) = 0.33, *p* = .861), attrition severity (*F*(4, 442) = 0.02, *p* = .982), and resorption severity (*F*(4, 437) = 0.08, *p* = .778) were not significantly associated with dentine lithium levels within the analyzed developmental window (-12 to +13 WSB) and therefore were not included as covariates in subsequent analyses.

Associations between dentine lithium levels and neuroimaging outcomes were evaluated using regional linear mixed-effects models across cortical thickness (68 regions-of-interest [ROIs]), cortical surface area (68 ROIs), and subcortical volume (16 ROIs). All analyses were restricted to participants who passed baseline T1-weighted MRI quality control procedures (*n* = 437; 97.5%). Cortical thickness models included fixed effects for age, sex, caregiver education, and whole-brain volume. Cortical surface area models substituted total cortical surface area as the modality-specific global covariate, whereas subcortical volume models adjusted for intracranial volume (*64*). Sex-by-age interaction terms were included to account for sex-specific developmental trajectories in brain maturation (*64*). Lithium-by-caregiver education interaction terms were included to test whether associations between dentine lithium levels and neurodevelopment varied across SES context (*46*). Lithium-by-WSB interaction terms were included to evaluate whether developmental timing of lithium exposure differentially related to neurodevelopmental outcomes across the exposure window. Random slopes for the lithium-by-WSB term were specified at the participant level to account for individual differences in temporally varying dentine lithium levels across WSB, and MRI device was modeled as a random intercept to account for scanner- related variability across ABCD acquisition sites. Neuroimaging models followed the general form:

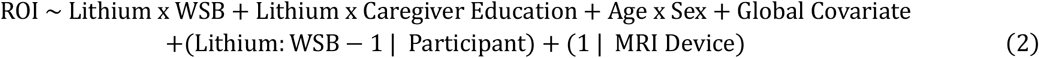

Separate sets of linear mixed-effects models were subsequently conducted to evaluate associations between dentine lithium levels and cognitive performance (NIH Toolbox) as well as emotional and behavioral problems (CBCL). Models included fixed effects for dentine lithium levels, WSB, caregiver education, age, sex, and the interactions lithium **×** WSB, lithium **×** caregiver education, and age **×** sex, with participant-level random slopes for the lithium **×** WSB term and a random intercept for assessment site. To reduce the burden of multiple testing, NIH Toolbox and CBCL analyses followed hierarchical testing frameworks progressing from broad composite measures to domain-specific subscales.

Follow-up ROI-to-outcome analyses examined whether lithium-associated neuroanatomical variation was associated with cognitive and behavioral outcomes. Participant-level dentine lithium levels were summarized using the mean of repeated log-transformed dentine lithium measurements across WSB. Separate linear regression models were then conducted for cortical thickness, cortical surface area, and subcortical volume ROIs, adjusting for participant-level mean dentine lithium levels, age, sex, caregiver education, assessment site, and modality-specific global neuroanatomical covariates.

Influential observations were identified across all linear mixed-effects models using participant-level Cook’s distance estimates derived with the influence.ME package (*65*). For each model, Cook’s distance was computed after refitting the model on complete-case data, and participants exceeding the 99^th^ percentile of the model-specific Cook’s distance distribution were classified as influential observations (*66*, *67*). Primary inferential results are reported from Cook’s-adjusted models, with corresponding non- adjusted models reported in Tables S2-S10.

To account for multiple testing, false discovery rate (FDR) correction was implemented using the Benjamini-Hochberg rank-based procedure (*68*) across neuroimaging and behavioral models following Cook’s distance-based exclusion of influential participant-level observations. Because neuroimaging analyses involved large numbers of regional outcomes, FDR correction was applied separately for cortical thickness, cortical surface area, and subcortical volume analyses using a conservative threshold of α < .005 to minimize type-I errors (*69*). Non-neuroimaging analyses, including cognitive and behavioral outcome models, were evaluated at α < .05. For each family of tests, *p*-values were ordered from smallest to largest and assigned a rank *i* among the total number of non-missing tests *m*. The Benjamini- Hochberg critical value was calculated as:

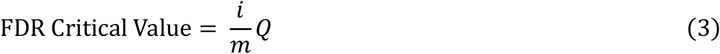

where *Q* represents the target FDR threshold. The largest rank *k* for which the raw *p*-value was less than or equal to its critical value was identified, and all tests with rank ≤ *k* were considered statistically significant.

Effect sizes were quantified using partial correlations derived from model *t*-statistics and residual degrees of freedom:

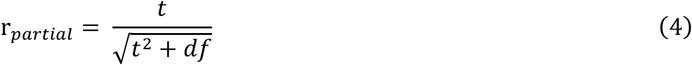

where *t* represents the regression *t*-statistic for the predictor of interest and *df* denotes the residual degrees of freedom from the fitted model (*70*, *71*). Ninety-five percent confidence intervals for partial correlations were estimated using the effectsize package (*72*). Raw *p*-values surviving FDR correction are reported throughout.

To translate the strongest lithium **×** caregiver-education interactions into original outcome units, we derived estimated marginal trends (simple slopes) for dentine lithium from the corresponding interaction models using the emmeans package (*73*). Trends were evaluated at three caregiver-education levels: the sample average (mean-centered 0) and the lowest (less than high school) and highest (graduate/professional degree) observed levels, to characterize both the overall association and its socioeconomic variation. Given the large sample, asymptotic degrees of freedom were used for all slope estimates and confidence intervals. Slopes and 95% confidence intervals, estimated on the mean- centered, log-transformed ^7^Li:^43^Ca ratio, were rescaled by the interquartile range (IQR) and standard deviation (SD) of dentine lithium to express effect sizes per IQR or per SD increase in early-life environmental lithium exposure.

To benchmark the overall (sample-average) association against the more established pediatric lead- exposure literature, we identified the corresponding mean-centered, log-scale slope at the sample- average caregiver-education level and rescaled it by ln(2) (= 0.693) to express the expected change in outcome per doubling of dentine lithium. Doubling of exposure is the metric conventionally used to express exposure-response slopes in log-linear lead models (*31*, *74*). Blood lead comparator estimates were taken directly, without re-derivation, from two published sources reporting effects on matching log- exposure scale: (1) the adjusted regression coefficient for log-transformed concurrent blood lead on full- scale IQ (β = -2.65 per unit increase in natural-log blood lead, 95% CI [-3.69, -1.61]), reported in Table 4 of the corrected pooled analysis by Lanphear et al. (*31*); and (2) the adjusted regression coefficient for ln- transformed blood lead concentration at age 8 years on externalizing problems (β = 3.1 per unit increase in natural-log blood lead, 95% CI [0.7, 5.4]), reported by Sears and colleagues (*32*). The age-8 estimate was selected both as the closest match to our cohort’s assessment age (9-10 years) and as the largest of six age-specific coefficients reported (ages 1-5 and 8), consistent with the original authors’ finding of heightened susceptibility at that age. Both comparator coefficients were rescaled by ln(2) identically to the dentine lithium slopes, yielding change per doubling of blood lead. As neither comparator study reports an SES-stratified coefficient on this scale, the lead-exposure benchmark is necessarily limited to the overall (marginal) lithium association; the education-stratified associations reported above and in Fig. S7 do not have a corresponding lead benchmark. Table S12 reports the full calculation, including source coefficients, confidence intervals, and rescaled estimates, to allow independent verification.

## Data Availability

Data used in the preparation of this article were obtained from the Adolescent Brain Cognitive Development (ABCD) Study, held in the NIH Brain Development Cohorts Data Sharing Platform (https://www.nbdc-datahub.org/; public data release 7.0, https://doi.org/10.82525/8f3w-5260). The ABCD data repository grows and changes over time. This is a multisite, longitudinal study designed to recruit more than 10,000 youth aged 9-10 and follow them over 10 years into early adulthood. The R code used for all analyses is available on the Open Science Framework (https://osf.io/u8v6e/overview?view_only=fcf2da63dc1e452495d30653bf78b82d). This study did not generate new materials.

## Acknowledgments

Funding

The ABCD Study^®^ is supported by the National Institutes of Health and additional federal partners under award numbers: U01DA041048, U01DA050989, U01DA051016, U01DA041022, U01DA051018, U01DA051037, U01DA050987, U01DA041174, U01DA041106, U01DA041117, U01DA041028, U01DA041134, U01DA050988, U01DA051039, U01DA041156, U01DA041025, U01DA041120, U01DA051038, U01DA041148, U01DA041093, U01DA041089, U24DA041123, U24DA041147. A full list of supporters is available at Federal Partners – ABCD Study (https://abcdstudy.org/about/federal-partners/).

Additional support for this work was made possible from: National Institute of Environmental Health Sciences grant R01-ES032295 (E.R.S.) National Institute of Environmental Health Sciences grant R01-ES031074 (E.R.S.) National Institute of Environmental Health Sciences grant R01-ES031247 (E.R.S.)

National Institute of Diabetes and Digestive and Kidney Diseases grant K01-DK135847 (S.A.) National Institute of Environmental Health Sciences grant P30-ES007048 (R.M.)

National Institute of Environmental Health Sciences grant P2C-ES033433 (R.M.) National Institute of Environmental Health Sciences grant R35-ES030435 (M.A.)

National Institute of Environmental Health Sciences grant R01-ES038499 (M.A., V.M., M.M.) National Institute of Environmental Health Sciences grant P30-ES023515 (M.A., V.M., M.M.)

## Author contributions

Conceptualization: M.K., A.T.M., V.M., M.A., E.R.S.

Methodology: M.K., A.T.M., S.A., R.M., M.M., V.M., M.A.

Investigation: A.T.M., E.K., M.M., M.A. Visualization: M.K., A.T.M. Supervision: M.A., E.R.S.

Writing – original draft: M.K., A.T.M., M.A., E.R.S.

Writing – review & editing: E.K., S.A., R.M., M.M., V.M.

ABCD Consortium investigators designed and implemented the study and/or provided data but did not necessarily participate in the analysis or writing of this report. This manuscript reflects the views of the authors and may not reflect the opinions or views of the NIH or ABCD Consortium investigators.

## Data, code, and materials availability

Data used in the preparation of this article were obtained from the Adolescent Brain Cognitive Development™ (ABCD) Study, held in the NIH Brain Development Cohorts Data Sharing Platform (https://www.nbdc-datahub.org/; public data release 7.0, https://doi.org/10.82525/8f3w-5260). The ABCD data repository grows and changes over time. This is a multisite, longitudinal study designed to recruit more than 10,000 youth aged 9-10 and follow them over 10 years into early adulthood. The R code used for all analyses is available on the Open Science Framework (https://osf.io/u8v6e/overview?view_only=fcf2da63dc1e452495d30653bf78b82d). This study did not generate new materials.

## Supplemental Results

### Cortical Thickness

Across cortical thickness ROIs, higher dentine lithium levels were consistently associated with greater cortical thickness. Of the 68 ROIs, 41 (60%) survived FDR correction and all significant associations were positive (Tables S5.1-S5.2). The strongest effects were observed in the right caudal middle frontal gyrus (*rp* = .11, 95% CI [.09, .13], *p* < .001), right pars opercularis of the inferior frontal gyrus (*rp* = .10, 95% CI [.09, .12], *p* < .001), right precuneus cortex (*rp* = .09, 95% CI [.07, .11], *p* < .001), left transverse temporal gyrus (*rp* = .09, 95% CI [.07, .11], *p* < .001), and left superior frontal gyrus (*rp* = .09, 95% CI [.07, .11], *p* < .001) (Fig. S3A, S4A).

Across 68 cortical thickness ROIs, 28 (41%) showed significant lithium **×** caregiver education interactions following FDR correction. Interactions were predominantly positive (71%), indicating that higher dentine lithium levels were associated with greater cortical thickness among youth whose caregiver reported higher educational attainment. Positive interactions were most prominent in frontal regions, with additional involvement of occipital and temporal cortices, including the bilateral rostral anterior cingulate (right: *rp* = .08, 95% CI [.06, .10], *p* < .001; left: *rp* = .13, 95% CI [.11, .15], *p* < .001), right cuneus (*rp* = .11, 95% CI [.09, .13], *p* < .001), and right superior frontal cortex (*rp* = .10, 95% CI [.08, .12], *p* < .001). A smaller, more distributed subset of regions (29%) showed negative interactions, indicating higher dentine lithium levels were associated with lower cortical thickness among youth with higher caregiver education levels. The strongest negative effect was observed in the left transverse temporal cortex (*rp* = -.08, 95% CI [-.10, - .06], *p* < .001) (Fig. 2A; Table S5.3).

### Surface Area

Across surface area ROIs, higher dentine lithium levels showed bidirectional and regionally heterogeneous associations. Of the 68 ROIs, 45 (66%) survived FDR correction, with a slight predominance of positive associations (58% positive; 42% negative) (Tables S6.1-S6.2). The strongest positive effects were observed in the left isthmus cingulate cortex (*rp* = .13, 95% CI [.11, .15], *p* < .001), left precuneus cortex (*rp* = .11, 95% CI [.09, .12], *p* < .001), left lateral orbitofrontal cortex (*rp* = .09, 95% CI [.07, .11], *p* < .001), and right lingual gyrus (*rp* = .09, 95% CI [.07, .11], *p* < .001). The strongest negative effect was observed in the right superior frontal gyrus (*rp* = -.10, 95% CI [-.11, -.08], *p* < .001) (Fig. S3B, S4B).

Across 68 cortical surface area ROIs, 44 (65%) showed significant lithium **×** caregiver education interactions following FDR correction. Interactions were equally distributed with positive (50%) and negative (50%) effects, indicating that associations between dentine lithium levels and cortical surface area varied heterogeneously as a function of caregiver education. Positive interactions were primarily observed in temporal and parietal regions, including the right middle temporal gyrus (*rp* = .11, 95% CI [.09, .13], *p* < .001), left supramarginal gyrus (*rp* = .10, 95% CI [.08, .12], *p* < .001), and left pericalcarine cortex (*rp* = .08, 95% CI [.06, .09], *p* < .001). A subset of regions showed negative interactions, concentrated in frontal and motor-related regions, including the left precentral gyrus (left: *rp* = -.16, 95% CI [-.17, -.14], *p* < .001) as well as the right inferior parietal cortex (*rp* = -.07, 95% CI [-.09, -.05], *p* < .001) (Fig. 2B; Table S6.3).

### Subcortical Volume

Across subcortical volume ROIs, higher dentine lithium levels were more often associated with smaller subcortical volumes. Of the 16 ROIs, 10 (63%) survived FDR correction, the majority of which were negative (80% negative; 20% positive) (Tables S7.1-S7.2). The strongest negative effects were observed in the right accumbens area (*rp* = -.12, 95% CI [-.14, -.10], *p* < .001), right ventral diencephalon (*rp* = -.07, 95% CI [-.09, -.06], *p* < .001), left putamen (*rp* = -.07, 95% CI [-.09, -.05], *p* < .001), and bilateral caudate (right: *rp* = -.06, 95% CI [-.08, -.05], *p* < .001; left: *rp* = -.07, 95% CI [-.09, -.05], *p* < .001) (Fig. S3C, S4C).

In contrast, positive associations were limited to the bilateral hippocampus (right: *rp* = .05, 95% CI [.03, .07], *p* < .001; left: *rp* = .04, 95% CI [.02, .05], *p* < .001).

Across 16 subcortical volume ROIs, 8 (50%) showed significant lithium **×** caregiver education interactions following FDR correction. All interactions were positive (100%), indicating that higher dentine lithium levels were associated with greater subcortical volumes among youth whose caregiver reported higher educational attainment. These effects were observed in limbic and striatal regions, including the bilateral hippocampus (right: *rp* = .10, 95% CI [.08, .12], *p* < .001; left: *rp* = .06, 95% CI [.04, .08], *p* < .001), left caudate (*rp* = .06, 95% CI [.04, .07], *p* < .001), and bilateral amygdala (right: *rp* = .05, 95% CI [.03, .07], *p* < .001; left: *rp* = .06, 95% CI [.04, .07], *p* < .001) (Fig. 2C; Table S7.3).

### Cognitive Performance (NIH Toolbox)

Higher dentine lithium levels were associated with lower overall cognitive performance (Tables S8.1-S8.2; Fig. S5). At the composite level, lithium was negatively associated with total cognition (*rp* = -.03, 95% CI [-.05, -.01], *p* = .008). At the domain level, higher dentine lithium levels were associated with lower crystallized composite scores (*rp* = -.06, 95% CI [-.08, -.04], *p* < .001), but not with fluid cognition (*rp* =.001, *p* = .892). At the task level, the crystallized effect was driven by Oral Reading Recognition (*rp* = -.14, 95% CI [-.16, -.11], *p* < .001), but not Picture Vocabulary (*rp* = -.001, *p* = .898). Each IQR increase in dentine lithium was associated with a 1.27-point decrease in Oral Reading Recognition (95% CI [-1.44, - 1.09], *p* < .001). Expressed per standard deviation of exposure, this corresponds to a 1.06-point decrease in Oral Reading Recognition (95% CI [-1.21, -0.92]). This association was substantially stronger among youth whose caregivers had less education and attenuated to near null among youth whose caregivers had more education (Fig. S7A; Table S11).

Across cognitive outcomes, several lithium **×** caregiver education interactions survived FDR correction. Significant interactions were observed for the total cognition composite (*rp* = .04, 95% CI [.02, .06], *p* < .001), crystallized cognition composite (*rp* = .06, 95% CI [.04, .08], *p* < .001) and Oral Reading Recognition (*rp* = .13, 95% CI [.11, .16], *p* < .001) (Fig. 3A). No interaction was observed for Picture Vocabulary (*rp* = .01, *p* = .228), suggesting heterogeneity across crystallized domains.

Although dentine lithium levels were not associated with fluid cognition at the main-effect level, significant lithium **×** caregiver education interactions emerged for the fluid cognition composite (*rp* = .03, 95% CI [.01, .05], *p* = .001), Dimensional Change Card Sort (*rp* = .11, 95% CI [.09, .13], *p* < .001), Picture Sequence Memory (*rp* = .06, 95% CI [.04, .08], *p* < .001), and List Sorting Working Memory (*rp* = .04, 95% CI [.02, .06], *p* < .001). No significant interactions were observed for Flanker Inhibitory Control (*rp* = -.01, *p* = .477) or Pattern Comparison Processing Speed (*rp* = .01, *p* = .586).

Across all significant models, interaction effects were positive, indicating that higher dentine lithium levels were associated with better cognitive performance among youth whose caregiver reported higher educational attainment (Table S8.3).

### Emotional and Behavioral Problems (Child Behavior Checklist)

Higher dentine lithium levels were associated with more behavioral problems (Tables S9.1-S9.2; Fig. S6). At the global level, lithium was positively associated with total problems (*rp* = .05, 95% CI [.03, .07], *p* < .001). At the domain level, higher dentine lithium levels were positively associated with greater externalizing problems (*rp* = .05, 95% CI [.03, .07], *p* < .001), but not internalizing problems (*rp* = .02, *p* = .082). Each IQR increase in dentine lithium was associated with a 1.39-point increase in externalizing problems (95% CI [1.11, 1.67], *p* < .001). Expressed per standard deviation of exposure, this corresponds to a 1.17-point increase in externalizing problems (95% CI [0.94, 1.41]). Again, this association was substantially stronger among youth whose caregivers had less education and attenuated to near null among youth whose caregivers had more education (Fig. S7B; Table S11).

Across emotional and behavioral outcomes, several lithium **×** caregiver education interactions survived FDR correction. Negative interactions were observed for total problems (*rp* = -.03, 95% CI [-.05, -.01], *p* = .001) and externalizing problems (*rp* = -.08, 95% CI [-.10, -.06], *p* < .001) (Fig. 3B), indicating that higher dentine lithium levels were associated with fewer behavioral problems among youth whose caregiver reported higher educational attainment. In contrast, a positive interaction was observed for internalizing problems (*rp* = .05, 95% CI [.03, .07], *p* < .001) (Fig. 3C), indicating that higher dentine lithium levels were associated with greater internalizing symptoms among youth whose caregiver reported higher educational attainment. Together, these findings suggest distinct moderation patterns across externalizing and internalizing domains (Table S9.3).

---

**Fig. S1.**
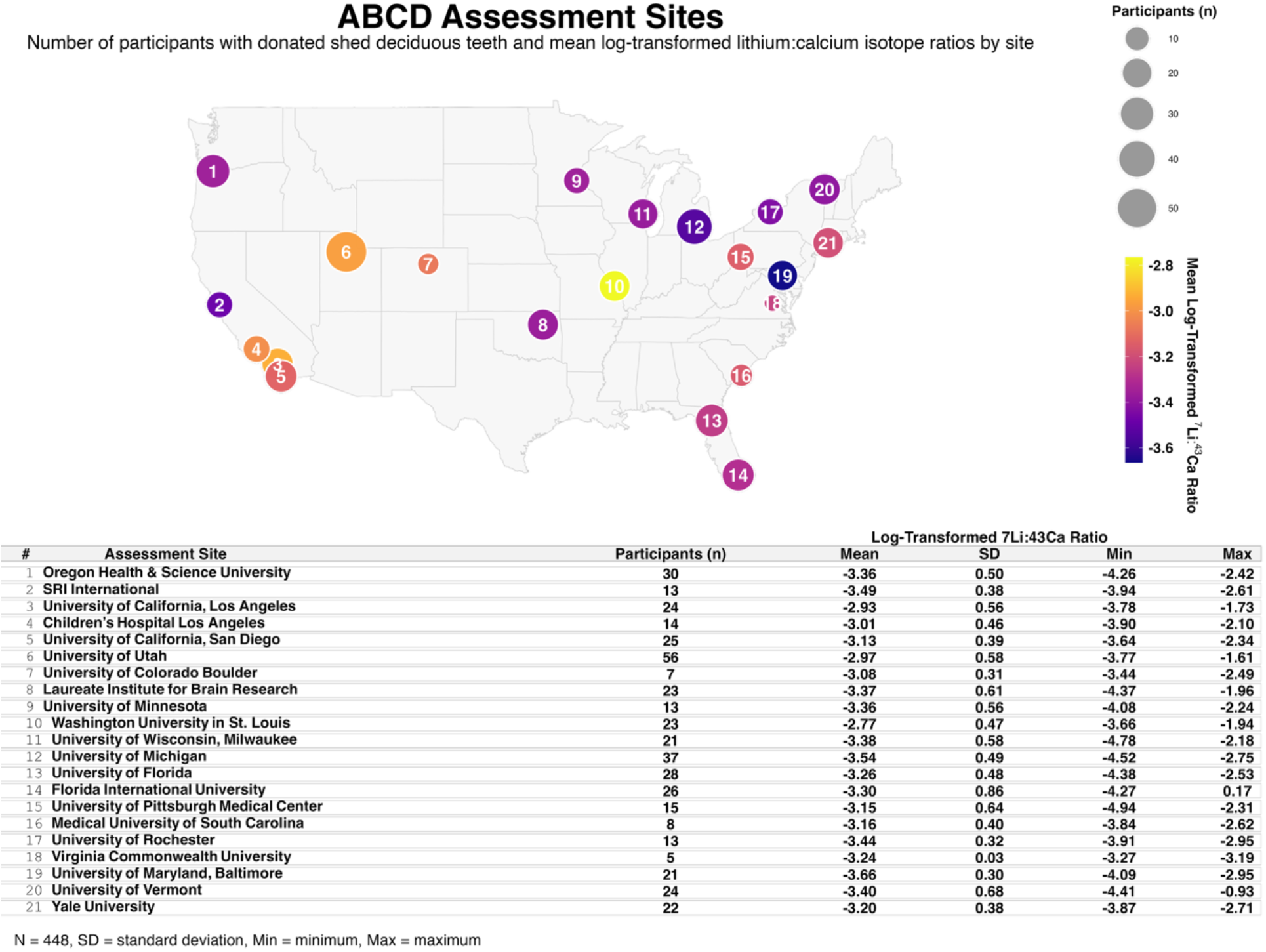
**ABCD assessment sites and site-level dentine lithium level distributions**. Geographic distribution of the 21 ABCD Study assessment sites contributing participants with donated shed deciduous teeth included in the dentine lithium biomarker subsample (*N* = 448). Circles indicate assessment site locations across the United States, with circle size representing the number of participants contributing teeth samples at each site. Circle color reflects the site-level mean log- transformed ^7^Li:^43^Ca ratio, with warmer colors indicating relatively higher dentine lithium levels and cooler colors indicating relatively lower levels. The accompanying table summarizes, for each assessment site, the number of participants, mean, standard deviation (SD), minimum, and maximum log-transformed ^7^Li:^43^Ca ratios.

**Fig. S2.**
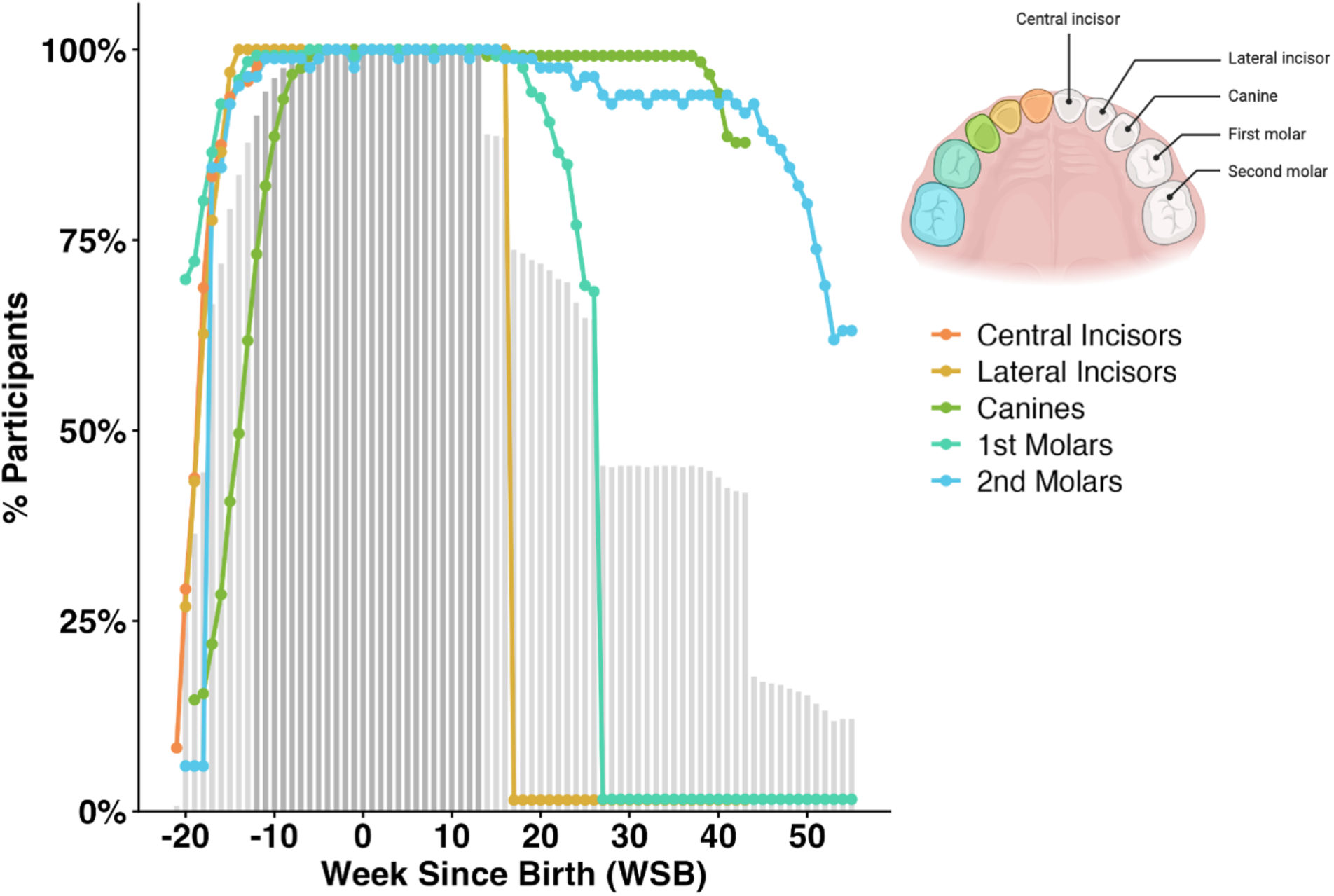
**Participant coverage across weeks since birth by deciduous tooth class**. Percentage of participants (y-axis) contributing early-life environmental lithium exposure data across weeks since birth (WSB; x-axis), stratified by deciduous tooth class. Colored lines indicate participant coverage for each tooth type (central incisors, lateral incisors, canines, first molars, and second molars), reflecting differences in developmental timing of tooth mineralization and crown formation. Grey bars represent overall participant coverage across all tooth classes combined, with the darker shaded regions highlighting the primary analytic window (-12 to +13). This interval was selected because it maximized participant coverage while minimizing missingness across individuals, with 90.8% of participants contributing exposure data during this period. Tooth schematic created with BioRender.com.

**Fig. S3.**
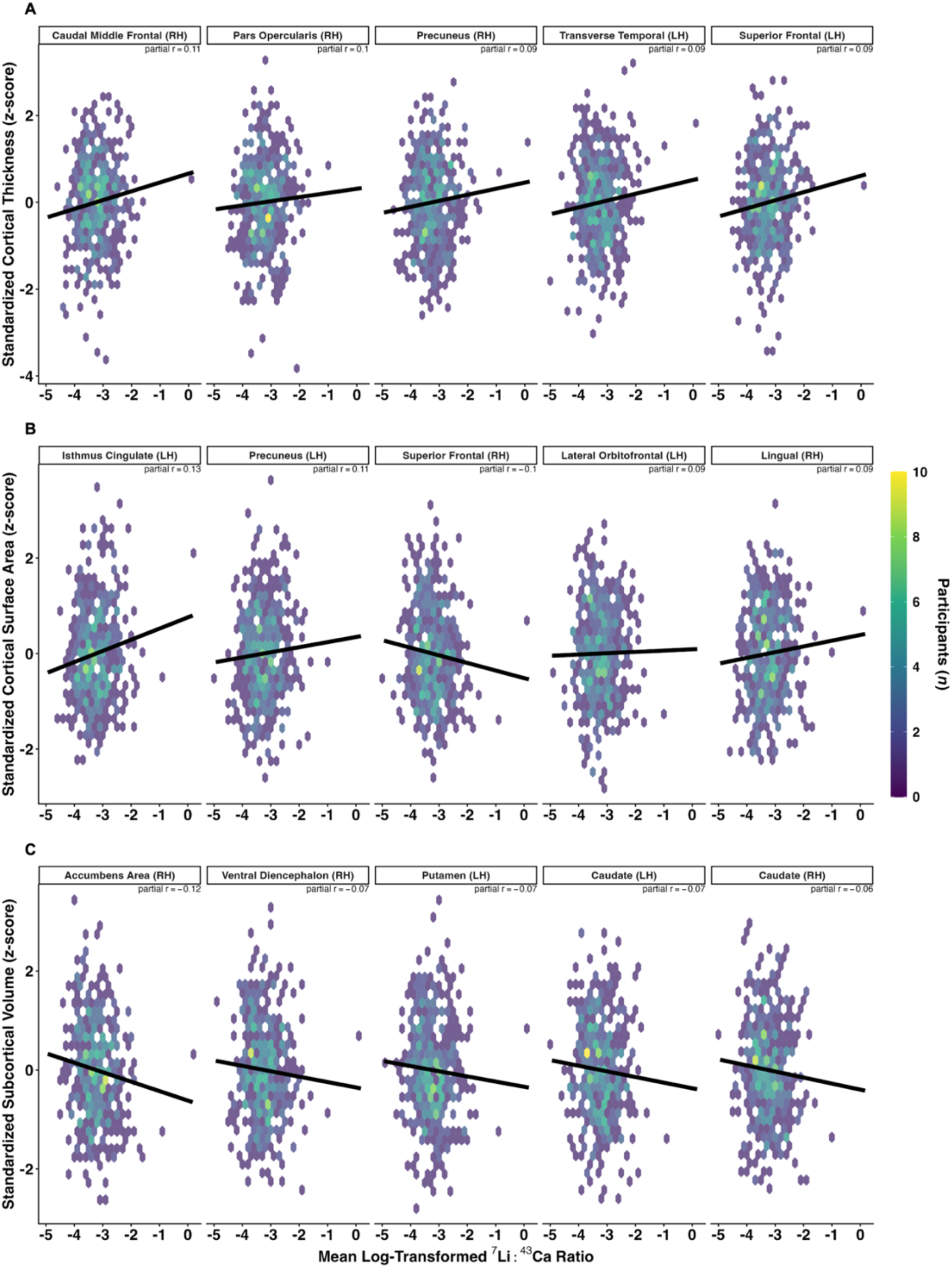
Associations between dentine lithium levels and the five strongest effects for cortical thickness, cortical surface area, and subcortical volume. Hexbin plots display associations between participants’ mean log-transformed ^7^Li:^43^Ca ratio (x-axis) and standardized (**A**) cortical thickness, (**B**) cortical surface area, and (**C**) subcortical volume measures (*z*-scores; y-axis) across regions showing the five strongest associations following FDR correction (Benjamini-Hochberg, α = .005). Regions were ranked by partial correlations (*rp*). Each hexagon represents the number of participants within bins of early-life environmental lithium exposure and brain structure measures, with brighter colors indicating greater density. Solid black lines denote linear regression fits estimated after exclusion of influential observations identified using Cook’s distance (99^th^ percentile threshold; *n* = 5 excluded). Partial correlation coefficients (*rp*), derived from covariate-adjusted linear mixed-effects models are displayed within each panel. Models were adjusted for age, sex, WSB (-12 to +13), caregiver education, and modality-specific global brain measures, including whole brain volume for cortical thickness analyses, total cortical surface area for cortical surface area analyses, and intracranial volume for subcortical volume analyses. The analytic sample after exclusion was *N* = 432, with the number of observations per region ranging from 11,120 to 11,126 due to repeated lithium measurements across timepoints. LH = left hemisphere; RH = right hemisphere.

**Fig. S4.**
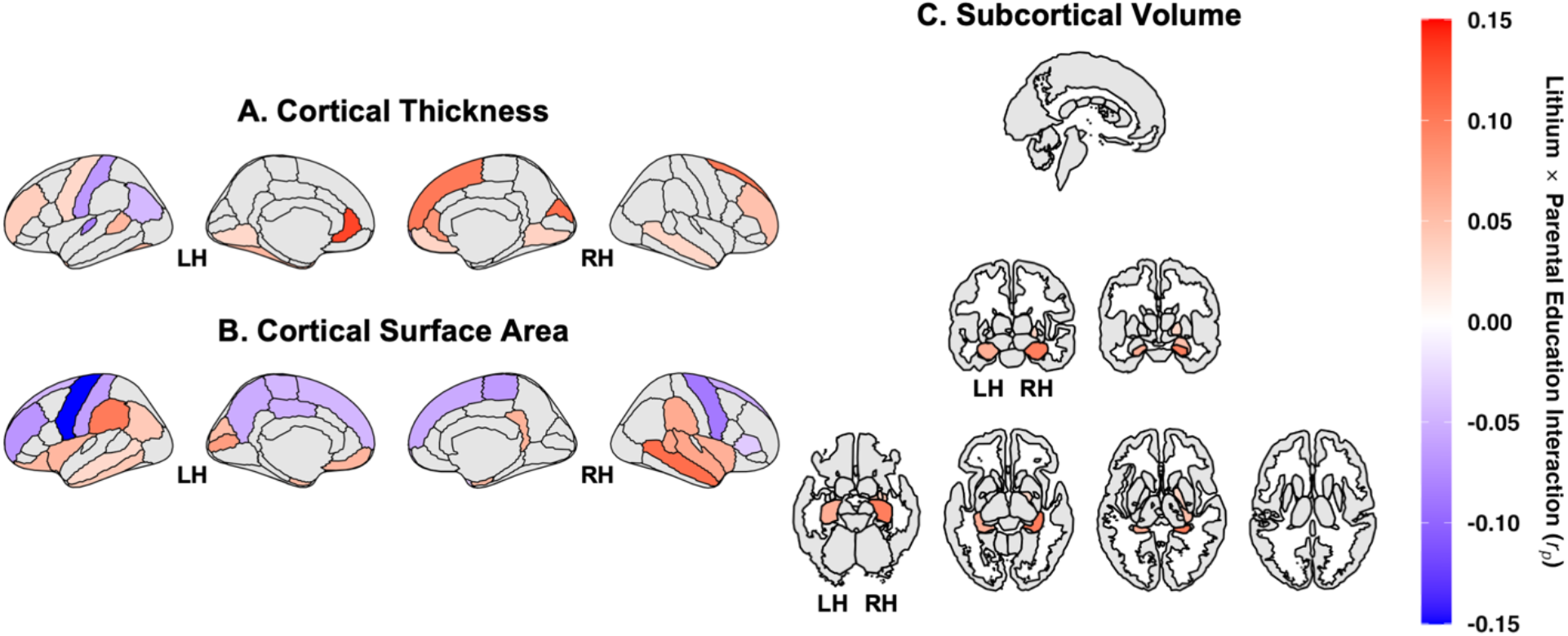
**Whole-brain lithium × caregiver education interaction effects on adolescent brain structure**. Surface-based and subcortical maps display partial correlations (*rp*) for the interaction between participants’ log-transformed ^7^Li:^43^Ca ratio and caregiver education in relation to (**A**) cortical thickness, (**B**) cortical surface area, and (**C**) subcortical volume. Cortical maps are shown for the left and right hemisphere (LH, RH) in lateral and medial views. Subcortical structures are visualized across multiple axial and coronal slices. Color values represent partial correlation coefficients (*rp*) derived from covariate- adjusted linear mixed-effects models, including age, sex, weeks since birth (-12 to +13), caregiver education, and modality-specific global brain measures. Warmer colors indicate positive associations, and cooler colors indicate negative associations. Only regions surviving FDR correction (Benjamini-Hochberg, α = .005) are displayed; non-significant regions are shown in grey. Analyses were conducted after exclusion of influential observations identified via Cook’s distance (99^th^ percentile threshold; *n* = 5 excluded), yielding a final sample of *N* = 432. The number of total observations per region ranged from 11,120 to 11,126 due to repeated measurements across timepoints.

**Fig. S5.**
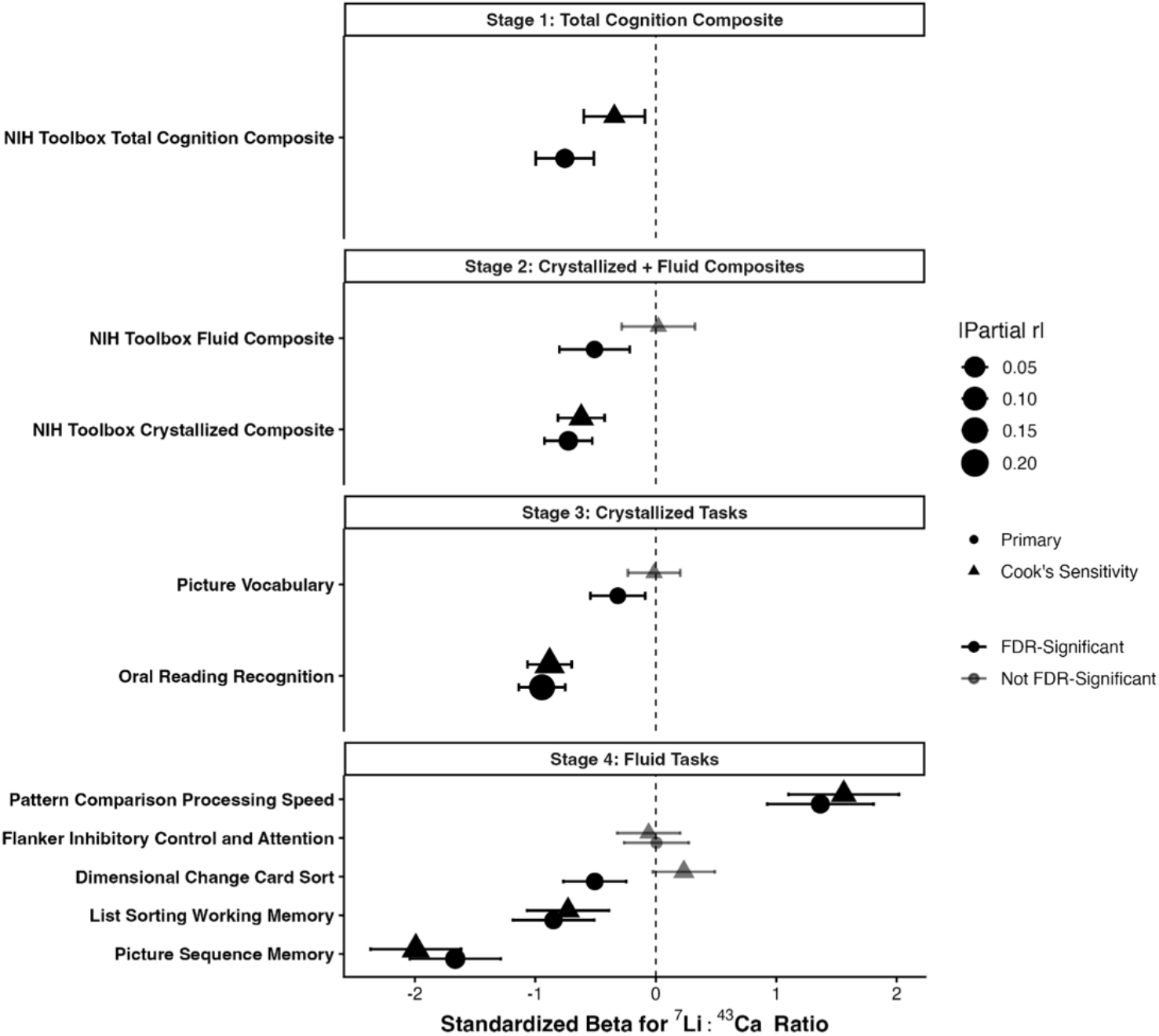
**Associations between dentine lithium levels and cognitive performance across hierarchical NIH Toolbox domains and tasks**. Forest plot showing standardized beta coefficients and partial correlations (|Partial r|) from linear mixed-effects models examining associations between log- transformed ^7^Li:^43^Ca ratios and cognitive performance on the NIH Toolbox Cognition Battery. Analyses followed a hierarchical testing framework progressing from broad composite measures (Stage 1) to domain-level composites (Stage 2) and individual task measures (Stages 3-4). Models included fixed effects for dentine lithium levels, weeks since birth (WSB), caregiver education, age, sex, and the lithium **×** WSB, lithium **×** caregiver education, and age **×** sex interaction terms, with participant-level random slopes for the lithium **×** WSB interaction and a random intercept for assessment site. Points indicate standardized beta estimates and horizontal error bars represent 95% confidence intervals. Circle size reflects the magnitude of the partial correlation coefficient. Black markers denote associations surviving FDR correction (Benjamini-Hochberg, α = .05), whereas grey markers indicate non-significant effects. Circles represent primary analyses and triangles represent Cook’s distance-adjusted sensitivity analyses (99^th^ percentile threshold). The vertical dashed line indicates the null effect (β = 0). Participant sample sizes ranged from *N* = 435-439 across NIH Toolbox outcomes depending on task-specific missingness, corresponding to 10,261-11,306 repeated observations across WSB.

**Fig. S6.**
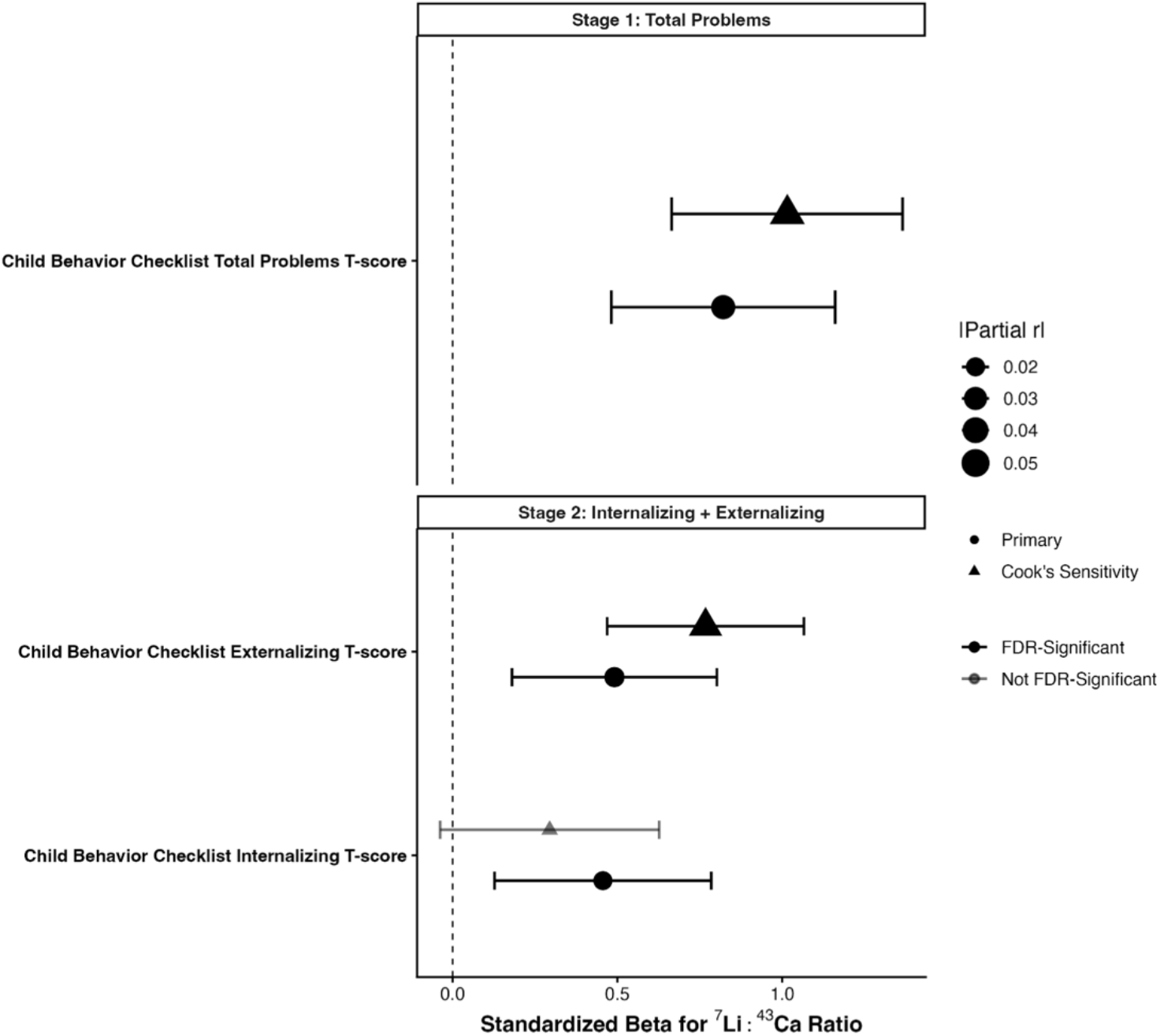
Associations between dentine lithium levels and emotional-behavioral problems across hierarchical Child Behavior Checklist domains. Forest plot showing standardized beta coefficients and partial correlations (|Partial r|) from linear mixed-effects models examining associations between log- transformed ^7^Li:^43^Ca ratios and emotional-behavioral problems on the Child Behavior Checklist (CBCL). Analyses followed a hierarchical testing framework progressing from broad total problem scores (Stage 1) to domain-level internalizing and externalizing problems (Stage 2). Models included fixed effects for dentine lithium levels, weeks since birth (WSB), caregiver education, age, sex, and the lithium **×** WSB, lithium **×** caregiver education, and age **×** sex interaction terms, with participant-level random slopes for the lithium **×** WSB interaction and a random intercept for assessment site. Points indicate standardized beta estimates and horizontal error bars represent 95% confidence intervals. Circle size reflects the magnitude of the partial correlation coefficient. Black markers denote associations surviving FDR correction (Benjamini-Hochberg, α = .05), whereas grey markers indicate non-significant effects. Circles represent primary analyses and triangles represent Cook’s distance-adjusted sensitivity analyses (99^th^ percentile threshold). The vertical dashed line indicates the null effect (β = 0). Participant sample was *N* = 443 across CBCL outcomes, corresponding to 11,405-11,406 repeated observations across WSB.

**Fig. S7.**
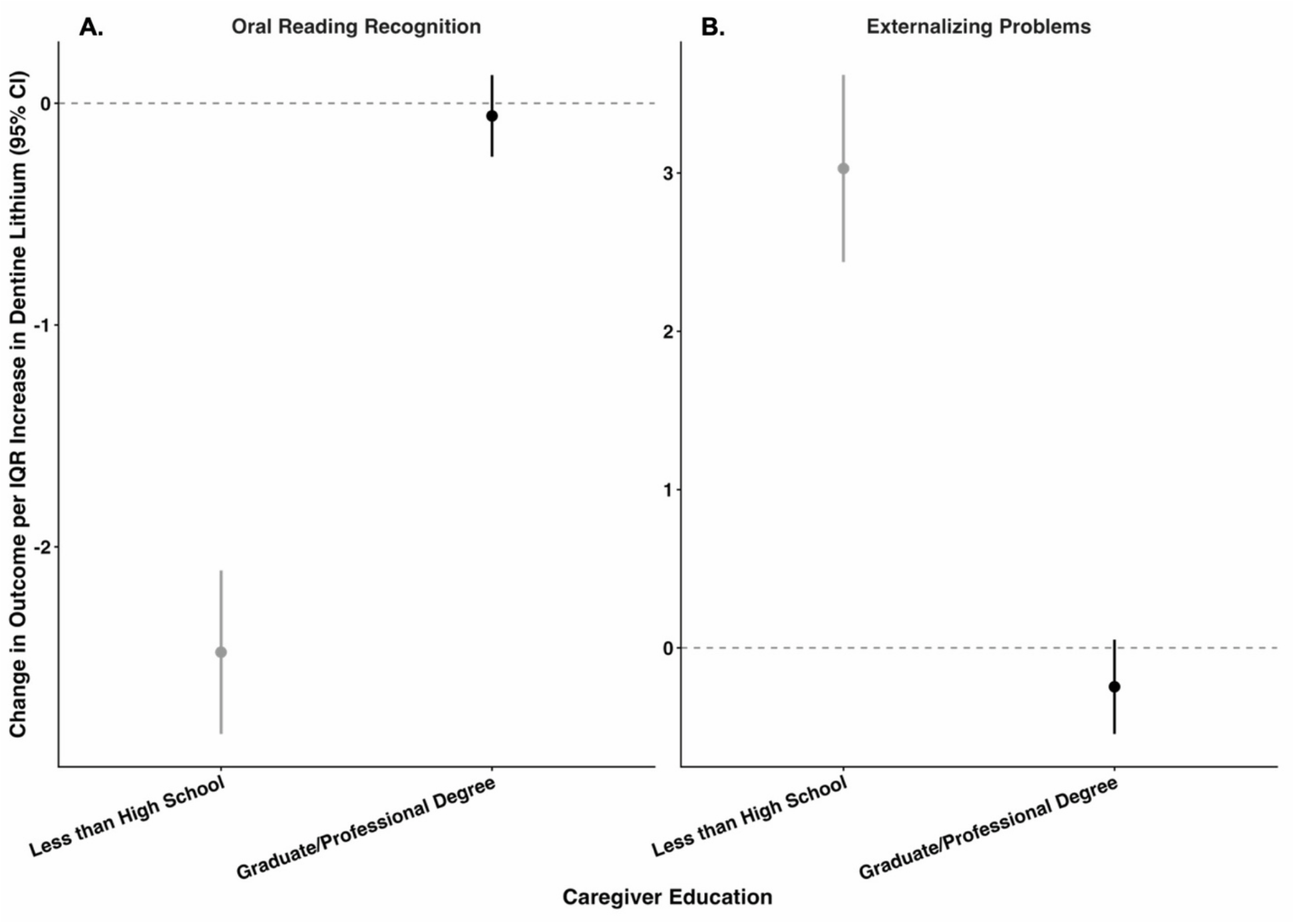
**Lithium × caregiver education simple-slope effect sizes for Oral Reading Recognition and externalizing problems**. Point-range plots display the estimated change in (**A**) Oral Reading Recognition (NIH Toolbox) and (**B**) externalizing problems (Child Behavior Checklist) per interquartile range (IQR) increase in participants’ log-transformed ^7^Li:^43^Ca ratio, separately for youth whose caregiver reported the lowest (less than high school; grey) versus highest (graduate/professional degree; black) level of educational attainment. Estimates reflect simple slopes (estimated marginal trends) derived from the same lithium **×** caregiver education linear mixed-effects interaction models reported in Tables S8.3 and S9.3, rescaled from the mean-centered model metric by the IQR of dentine lithium to express effect sizes in original units (uncorrected standard score points for Oral Reading Recognition; *T*-score points for externalizing problems). Points indicate the estimated change in outcome per IQR increase in dentine lithium, and vertical error bars represent 95% confidence intervals (calculated using asymptotic degrees of freedom). The horizontal dashed line indicates the null effect (no change in outcome). Analyses were conducted after exclusion of influential observations identified via Cook’s distance (99^th^ percentile threshold; *n* = 5 excluded for each outcome), yielding *N* = 439 participants and 11,303 repeated observations for Oral Reading Recognition and *N* = 443 participants and 11,405 repeated observations for externalizing problems.

**Table S1.**
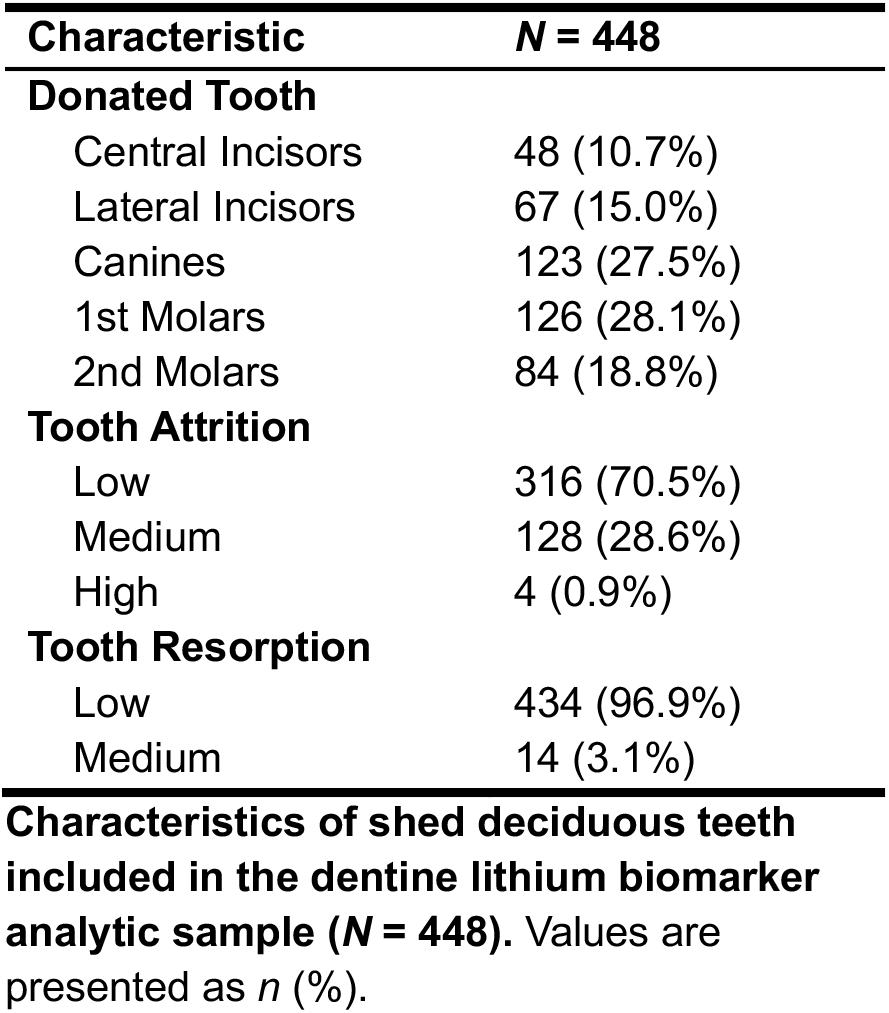
Characteristics of shed deciduous teeth included in the dentine lithium biomarker analytic sample (*n* = 448)

**Table S2.1.** Association between caregiver education and dentine lithium levels (unadjusted)

|  | ***r<sub>p</sub>*** | ***t (df)*** | ***p*** | **95% CI** |
|---|---|---|---|---|
| Intercept | -.04 | -0.20 (23) | .842 | [-.41, .35] |
| Sex | -.07 | -1.57 (438) | .116 | [-.17, .02] |
| WSB | .17 | 17.78 (11088) | < .001 | [.15, .18] |
| Caregiver Education | -.14 | -3.01 (448) | .003 | [-.23, -.05] |

**Table S2.2.** Association between caregiver education and dentine lithium levels (cook’s distance-adjusted)

|  | ***r<sub>p</sub>*** | ***t (df)*** | ***p*** | **95% CI** |
|---|---|---|---|---|
| Intercept | -.07 | -0.33 (23) | .741 | [-.43, .32] |
| Sex | -.06 | -1.26 (432) | .210 | [-.15, .03] |
| WSB | .17 | 18.01 (10966) | < .001 | [.15, .19] |
| Caregiver Education | -.16 | -3.48 (443) | < .001 | [-.25, -.07] |

**Table S3.1.** Association between area deprivation index and dentine lithium levels (unadjusted)

|  | ***r<sub>p</sub>*** | ***t (df)*** | ***p*** | ***95% CI*** |
|---|---|---|---|---|
| Intercept | -.06 | -0.29 (22) | .773 | [-.43, .34] |
| Sex | -.04 | -0.89 (430) | .373 | [-.14, .05] |
| WSB | .16 | 16.82 (10938) | < .001 | [.14, .18] |
| ADI | .11 | 1.84 (296) | .067 | [-.01, .22] |

**Table S3.2.** Association between area deprivation index and dentine lithium levels (cook’s distance-adjusted)

|  | ***r<sub>p</sub>*** | ***t (df)*** | ***p*** | **95% CI** |
|---|---|---|---|---|
| Intercept | -.09 | -0.41 (22) | .688 | [-.45, .31] |
| Sex | -.03 | -0.66 (426) | .509 | [-.13, .06] |
| WSB | .17 | 17.43 (10813) | < .001 | [.15, .18] |
| ADI | .12 | 2.10 (298) | .036 | [.01, .23] |

**Table S4.1.**
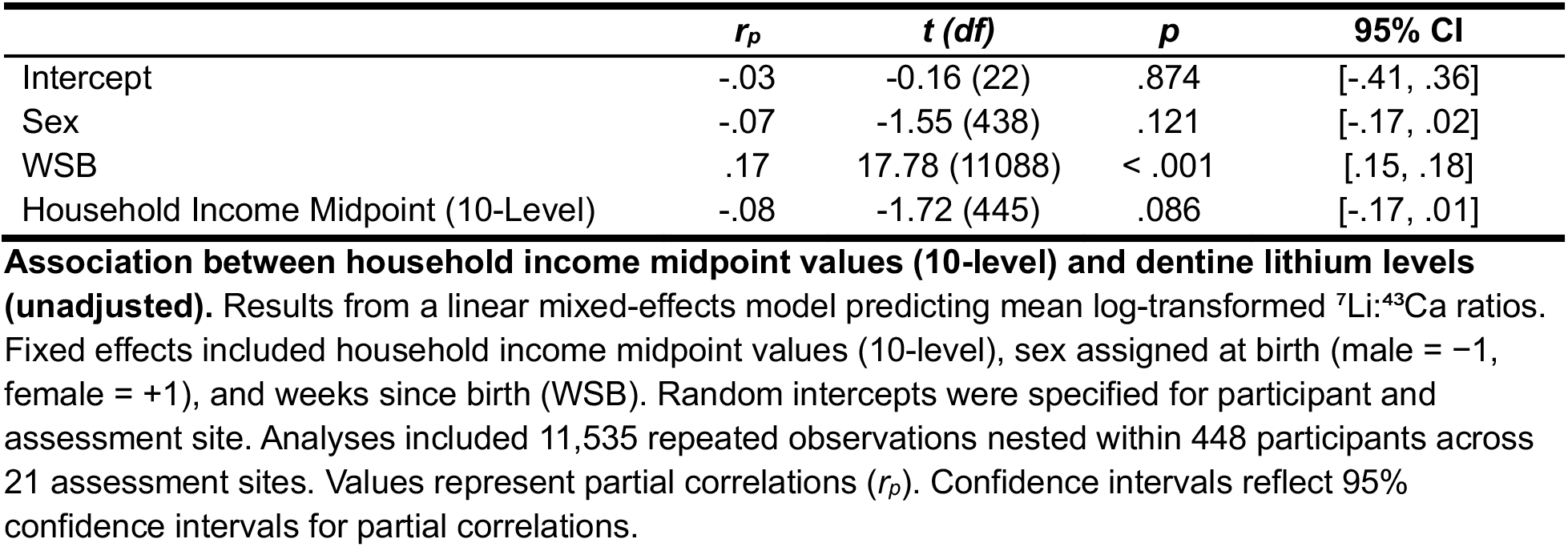
Association between household income midpoint values (10-level) and dentine lithium levels (unadjusted)

|  | ***r<sub>p</sub>*** | ***t (df)*** | ***p*** | **95% CI** |
|---|---|---|---|---|
| Intercept | -.03 | -0.16 (22) | .874 | [-.41, .36] |
| Sex | -.07 | -1.55 (438) | .121 | [-.17, .02] |
| WSB | .17 | 17.78 (11088) | < .001 | [.15, .18] |
| Household Income Midpoint (10-Level) | -.08 | -1.72 (445) | .086 | [-.17, .01] |

**Table S4.2.** Association between household income midpoint values (10-level) and dentine lithium levels (cook’s distance-adjusted) Due to the large number and size of regional association tables, supplemental Tables S5-S12 are provided in a separate Excel workbook titled *supplemental_tables_s5-s12_lithium-adolescent- neurodevelopment_11-02-2026.xlsx*. The workbook contains individual worksheets corresponding to each supplemental table and includes regional cortical thickness, cortical surface area, subcortical volume, NIH Toolbox, CBCL, and brain-behavior association, and lithium **×** caregiver education effect-size results.

|  | ***r<sub>p</sub>*** | ***t (df)*** | ***p*** | **95% CI** |
|---|---|---|---|---|
| Intercept | -.08 | -0.40 (22) | .691 | [-.45, .31] |
| Sex | -.05 | -0.97 (432) | .331 | [-.14, .05] |
| WSB | .17 | 18.04 (10963) | < .001 | [.15, .19] |
| Household Income Midpoint (10-Level) | -.07 | -1.44 (442) | .151 | [-.16, .02] |

## Supplemental Materials and Methods

All analyses were conducted in R version 4.5.2 (*1*), using the R packages for mixed-effects modeling, effect-size estimation, influential-case diagnostics, data processing, visualization, and table generation (*2–27*).

